# Model-informed target product profiles of long acting-injectables for use as seasonal malaria prevention

**DOI:** 10.1101/2021.07.05.21250483

**Authors:** Lydia Burgert, Theresa Reiker, Monica Golumbeanu, Jörg J. Möhrle, Melissa A. Penny

## Abstract

Seasonal malaria chemoprevention (SMC) has proven highly efficacious in reducing malaria incidence. However, the continued success of SMC is threatened by the spread of resistance against one of its main preventive ingredients, Sulfadoxine-Pyrimethamine(SP), operational challenges in delivery, and incomplete adherence to the regimens. Via a simulation study with an individual-based model of malaria dynamics, we provide quantitative evidence to assess long-acting injectables (LAIs) as potential alternatives to SMC. We explored the predicted impact of a range of novel preventive LAIs as a seasonal prevention tool in children aged three months to five years old during late-stage clinical trials and at implementation. LAIs were co-administered with a blood-stage clearing drug once at the beginning of the transmission season. We found the establishment of non-inferiority of LAIs to standard 3 or 4 rounds of SMC with SP-amodiaquine was challenging in clinical trial stages due to high intervention deployment coverage. However, our analysis of implementation settings where the achievable SMC coverage was much lower, LAIs with fewer visits per season are potential suitable replacements to SMC. Suitability as a replacement with higher impact is possible if the duration of protection of LAIs covered the duration of the transmission season. Furthermore, optimizing LAIs coverage and protective efficacy half-life via simulation analysis in settings with an SMC coverage of 60% revealed important trade-offs between protective efficacy decay and deployment coverage. Our analysis additionally highlights that for seasonal deployment for LAIs, it will be necessary to investigate the protective efficacy decay as early as possible during clinical development to ensure a well-informed candidate selection process.

## 2. Background

Children carry the majority of the global malaria burden, with an estimated 67% of all malaria-related deaths (272 000 in 2019) occurring in those under 5 years of age ^1^. In addition to effective and timely treatment, prevention through vector control or drug-based prophylaxis has proven to be an effective approach, reducing incidence and mortality ^2^. Especially in seasonal malaria transmission settings, where malaria transmission is linked to the rainy months, well-timed preventive malaria interventions that protect from infection during the transmission months can ease a substantial amount of malaria burden^1^. The WHO has recommended seasonal malaria chemoprevention (SMC) with monthly Sulfadoxine-Pyrimethamine+Amodiaquine (referred to as SMC-SP+AQ) for children aged between 3 months and 5 years during the malaria transmission season since 2012 ^3^. SP+AQ provides a two-stage effect: while AQ clears existing blood-stage infections, the long clearance half-life of SP prevents new infections. The impact of SMC in seasonal settings has been widely demonstrated, achieving a protective efficacy of roughly 80% against clinical episodes in a trial in Burkina Faso ^4^, a reduction in incidence by 60% in routine implementation in Senegal (80% deployment coverage of all eligible children) ^5^, and a reduction in the proportion of positive tests by 44% in routine implementation in Mali ^6^.

Despite its potential, poor adherence and the spread of drug resistance limit the effectiveness of SMC. Additionally, the monthly delivery of SMC-SP+AQ (one day of SP and three days of AQ) throughout the transmission season is relatively expensive, due to human resources and especially due to operational constraints during the rainy season ^7^. Consequently, in 2019, only 62% of children living in SMC-eligible areas in the Sahel subregion received SMC ^1^. Throughout the transmission season, coverage subsequently decreased by 6% in Guinea ^8^ and 20% in Mali ^9^. Investigation of the adherence to the 3x AQ regimen within one treatment course in a clinical trial in Niger showed that only 20% of children received the full regimen^10^. Additionally, the spread of resistance markers against SP was reported with increasing SMC deployment ^11, 12^, impacting the eligibility of regions for SMC ^13^ as well as the protective efficacy after implementation ^14^.

The need for alternative prevention tools that simplify deployment and possess a reduced risk of resistance is pressing. In the absence of an effective vaccine, long-acting injectables (LAIs) with an anti-infective effect could provide an alternative seasonal prevention tool by simplifying the deployment and reducing the risk of resistance through decreased selection pressure for SP resistance ^15^. Current candidate LAIs include small molecule drugs ^16,17^ or monoclonal antibodies (mAbs) ^18,19^ that target the sporozoite stage or liver stage of the malaria parasite, thereby serving as anti-infectives. The successful development of a LAI entails the definition of appropriate product profiles and use cases which are specified in Target Product Profiles (TPPs). Precisely, these specifications include LAI efficacy and safety prerequisites as well as delivery modalities ^15^. TPPs are living documents and therefore continuously updated as new evidence for product requirements becomes available.

To justify the implementation of LAIs under the use case of seasonal malaria prevention, non-inferiority to the existing intervention of SMC-SP+AQ must be met ^4^. For new tools with new modes of action and/or deployment modalities, proving non-inferiority to the standard of care is a crucial step and is usually established in non-inferiority clinical trials conducted at late stages of tool development. Currently, it is not yet known what clinical studies will be required for LAIs. In absence of efficacy data on LAIs, it is at the current stage impossible to obtain insights about the circumstances in which LAIs have the potential to be non-inferior. *In silico* modelling and simulation approaches of malaria transmission and control, allow the quantification of the impact of varying tool specifications in relation to varying operational and setting constraints which would not be feasible in real life clinical trials. They thus allow for the exploration of the potential to meet non-inferiority criteria. In the absence of non-inferiority evidence at the early stages of development, modelling and simulation approaches therefore provide a quantitative basis to further inform decision making. They guide tool development from the early stages by predicting potential public health impact (Fig. 1a) ^20^ and understanding non-inferiority criteria prior to clinical trial planning.

**Figure 1:**
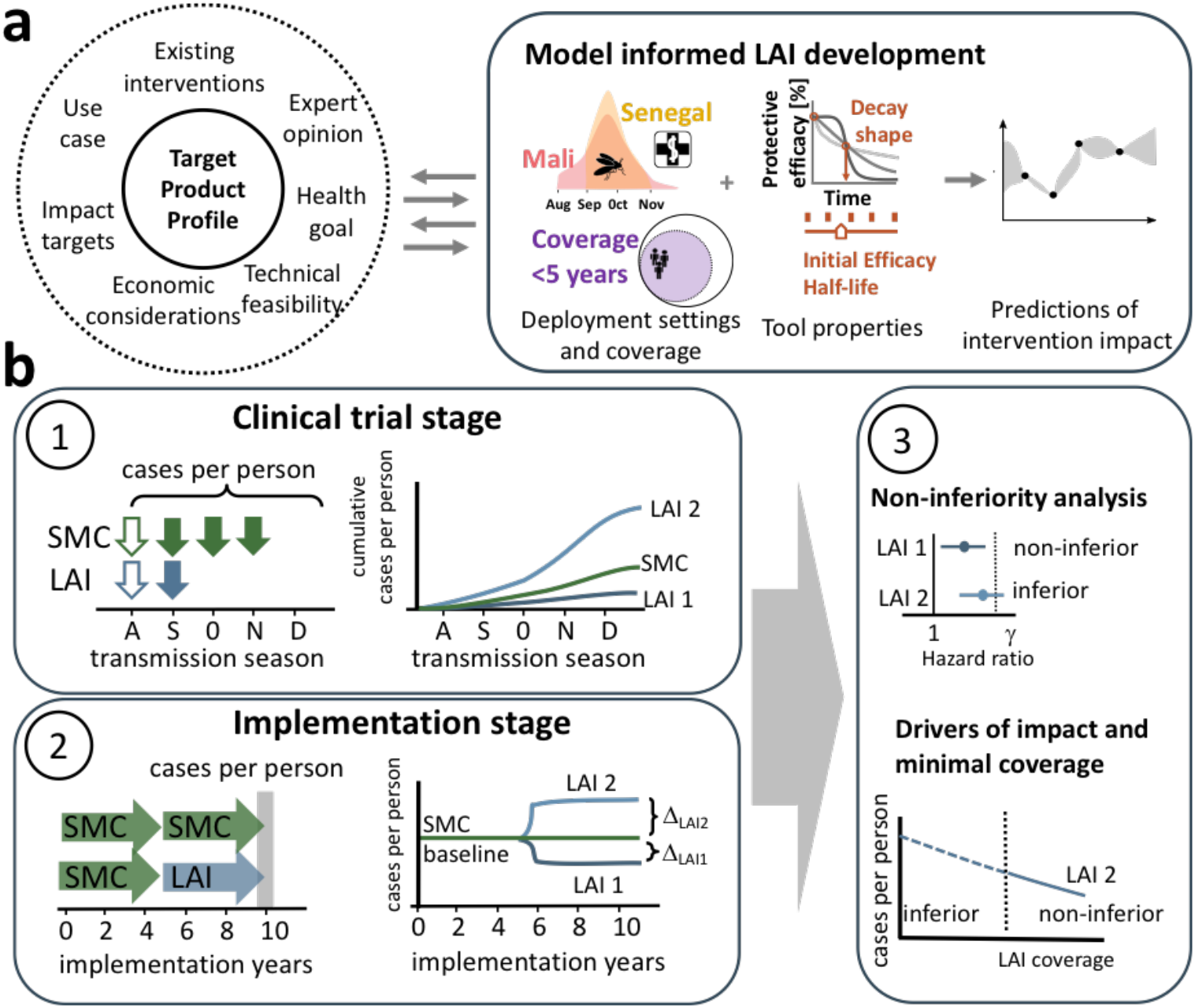
Workflow to assess the target product profile of LAIs. **(a)** In an iterative exchange between various stakeholders, definition of TPPs is informed by results from modelling approaches. Simulation of predefined scenarios with *OpenMalaria*, an individual based model of malaria dynamics, allows to estimate the impact (incidence reduction) of LAIs in the context of deployment setting details (access to healthcare and seasonality), deployment coverage of the target population and the tool properties (initial protective efficacy against infection, protective efficacy half-life and decay of protective efficacy). The resulting evaluation of LAI impact is communicated and discussed with stakeholders to refine the analysis as LAIs are developed. **(b)** The analysis in the clinical trial and implementation stage are illustrated on the example of two hypothetical LAIs with different efficacy profiles (denoted LAI 1 and 2). (1) In clinical trial stages, the minimum essential properties to reach a certain health goal are evaluated in a two-arm clinical trial. SMC-SP+AQ is administered (green arrows) three (as in Senegal, filled arrows) or four (as in Mali, filled and unfilled arrows) times over the transmission season. LAIs are administered once at the beginning of the transmission season (blue filled arrow for Senegal and blue empty arrow for Mali). The cumulative cases per person over the trial period are tracked. (2) In implementation stages, SMC-SP+AQ is replaced with LAIs after five years of implementation. Impact is assessed in the last implementation year (grey bar) and compared to the baseline of SMC-SP+AQ implementation. Coverages of SMC-SP+AQ and LAIs are independent from each other. (3) Upper panel: the tool properties influencing the establishment of non-inferiority of LAIs to SMC are investigated in the clinical trial stage. Lower panel: at a fixed SMC deployment coverage, the minimum coverage of LAI deployment required to establish non-inferiority is identified.

Here, we investigate the potential public-health impact for various use cases of LAIs by conducting an *in silico* simulation analysis examining product properties and operational modalities supporting LAI implementation as a seasonal malaria prevention tool. Accordingly, in the simulated scenarios, LAIs were delivered to children under five once at the beginning of the transmission season with an antimalarial in settings currently approved for SMC deployment. Their protective effect was then compared to SMC-SP+AQ administered three or four times per season in monthly intervals. By combining disease modelling and simulation experiments with machine learning approaches, we efficiently explored the large space of possible parameter values describing intervention and transmission setting characteristics and analyzed trade-offs between tool characteristics and operational constraints in a variety of transmission settings ^20^. We conducted our malaria transmission simulations using *OpenMalaria*, an established individual-based stochastic simulation platform of malaria transmission and control ^21 22^. Based on this approach, we defined a quantitative framework to assess the influence of tool properties and operational constraints on the impact of SMC and LAIs. Using this framework, we investigated two malaria transmission settings based on the malaria transmission profile in Senegal and Mali and assessed public health impact for a plethora of different tool properties, deployment coverages and over multiple transmission intensities. Our analysis was carried out along the clinical development pathway from late clinical trials through to implementation of future LAIs as an SMC replacement. By understanding the main drivers of impact to reach a pre-defined health goal in implementation stages and under operational constraints, we provide an assessment of endpoints in clinical trials of newly developed LAIs and identify efficacy requirements for further development.

## 3. Materials and Methods

The impact of novel anti-infective LAIs depends on the tool properties defining their efficacy profile, as well as on the operational constraints and the respective underlying malaria epidemiology, that influence tool suitability for implementation in a given setting (Fig. 1a). We investigated the influence of tool properties over a large range of specified protective efficacies, as well as operational considerations (coverage of children) in several implementation settings varying in seasonality and access to healthcare. The drivers of predicted impact for preventive LAIs were analysed along their clinical development from clinical trials to implementation (Fig. 1b) and compared to current standard of care (SMC-SP+AQ). Accordingly, we defined two analysis stages: in the *clinical trial stage*, we investigated the maximum incidence reduction and the ability to establish non-inferiority to SMC-SP+AQ over one malaria season (Fig. 1 b, panel 1). In the *implementation stage*, we replaced SMC-SP+AQ with LAIs after five years of implementation at varying coverages, and we inferred the minimal LAI coverage at which LAIs are equivalent (non-inferiority) in public-health impact (incidence reduction) to a continued implementation of SMC-SP+AQ (Fig. 1b, panel 2).

We adapted a previously developed framework to inform TPPs of new interventions against infectious diseases ^20^. First, a set of simulated scenarios was defined. These were characterized by the delivery modality, tool specifications, and settings in which a concrete health target was analysed (in our case, incidence reduction). Second, a set of disease scenarios were simulated randomly over the entire parameter space to evaluate the health outcomes. The resulting database of simulations was used to train a Gaussian process emulator (GP), that predicts the health outcome given a set of input parameters. Third, the emulator was employed to perform sensitivity analysis and optimisation of tool properties with respect to health outcomes. This analysis allowed us to define the optimal product characteristics of a LAI that maximises the chance of achieving a desired health goal.

### Malaria disease transmission model

We explored the dynamics of a preventive LAI against malaria using *OpenMalaria*, a stochastic, individual-based simulator of malaria infection in humans linked to a deterministic model of the mosquito feeding cycle ^23,24^. *OpenMalaria* accounts for heterogeneity within a human population on multiple levels including host exposure, susceptibility and immune response ^21,25,26^. The model allows the investigation of interventions against malaria at multiple points in the malaria life cycle (e.g. vaccines ^22^, insecticide treated bed-nets, and reactive case detection ^27^) while monitoring a variety of health outcomes (e.g. prevalence, incidence, mortality) ^28^.

### Simulated disease scenarios

Using *OpenMalaria*, we simulated a range of transmission settings (Fig. S1) and assumptions for the implementation of SMC and LAIs as a seasonal infection prevention intervention replacing existing prevention with SMC-SP+AQ. These assumptions are with regards to the properties of the setting (seasonality and intensity of transmission), health system (access to care and treatment of clinical cases), new and replacement intervention (Table 1).

**Table 1:**
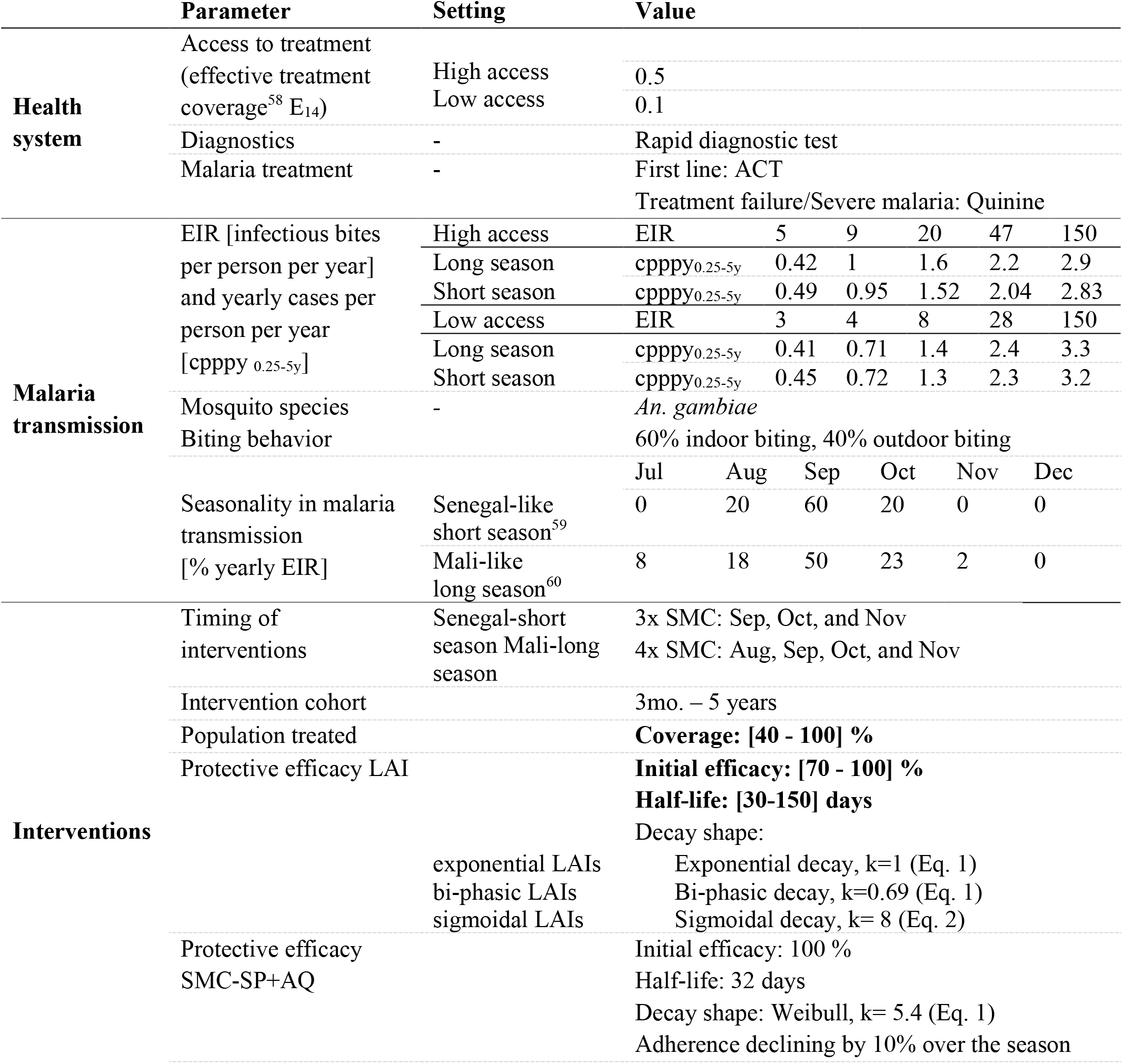
Summary of simulation set-up used for the implementation experiments. The simulated transmission settings were defined using a factorial design covering all possible combinations of discrete health system and vector specifications. The parameters defining the efficacy and delivery profiles of LAIs (highlighted in bold in the third column) were sampled within the defined parameter space using Latin Hypercube Sampling and simulated for each combination of settings. The effective coverage E_14_ describes the probability that effective malaria treatment will occur within a 14-day period since symptoms onset. Additional information on simulated transmission intensity can be found in the Supplement (Fig. S1 and Table S1).

The intervention age-group consisted of children between 3 months and 5 years of age (accounting for ca. 16% of the total population). The intervention age-group was chosen according to WHO recommendations ^3^, although some countries have implemented SMC for children up to 10 years old ^29^. We assumed a total population of 10,000 individuals to capture transmission settings within health facility catchment areas with an age-structure that represents a realistic age-distribution for African malaria-endemic settings ^30^. Access to treatment, defined as the percentage of the whole population who seek care for a symptomatic malaria episode, was chosen to represent settings with low or high health systems strength. The probability of symptomatic cases to receive effective treatment within two weeks from the onset of symptoms (E_14_) was set to 10% in low access to health-care settings and 50% in high access to health-care settings ^31^. The malaria seasonality profile, mosquito species and timing of interventions were parameterised to reflect those of Mali or Senegal, two countries in the Sahel region where SMC is implemented and clinical trials for malaria interventions are conducted frequently. In Mali, the malaria season is longer, starting in August and lasting until November (*long season*), and SMC is generally four monthly doses. In contrast, the malaria season in Senegal is only three months long, with a sharper profile (*short season*) and SMC is three doses one month apart (Fig. 1a, Table 1, Figure S2). Malaria prevalence in Senegal is generally low, with the highest *P. falciparum* prevalence in 2-10 years old (PfPR_2-10y_) in the southern regions being around 8%. However, the PfPR_2-10y_ in Mali is around 80% in the south of Mali but very low in the North ^32^.

To develop a broader understanding of the influence of transmission intensity on LAI impact, we simulated a range of initial incidence settings capturing the transmission heterogeneity of these two malaria-endemic countries (Figure S1, Table S1). The simulated transmission intensity of each setting was defined by the entomological inoculation rate (EIR) modelled as the average annual number of infectious bites received by an individual, and the corresponding simulated PfPR_2-10y_ (Table S1, Fig. S1). The protective efficacy of SMC-SP+AQ was implemented as being fully effective (no prevalent resistance against SP) or reduced duration of protection (prevalent resistance against SP).

Over all simulation experiments, the input parameter space for the protective efficacy and its decay, and the intervention coverage are as defined in Table 1. Parameters were randomly sampled using Latin hypercube sampling ^33^ by drawing 1500 parameter sets for each setting, with 5 stochastic realizations (simulation replicates) for each point. All simulations were performed using *OpenMalaria* version 38. The source code and comprehensive documentation for *OpenMalaria*, including a detailed model of demography, transmission dynamics and interventions is available at ^34^.

### Delivery: clinical trial and implementation assumptions

In our study, LAIs were implemented as anti-infective entities in the form of mAbs or small molecule drugs, that prevent the development of blood-stage malaria through action on malaria parasites in sporozoite or liver stages. We assumed administration once at the beginning of the transmission season in combination with an artemisinin combination therapy that cures prevalent blood-stage malaria infections with a 100% cure rate. SMC-SP+AQ was administered monthly: three times per season in Senegal like settings *(short season*) and four times per season in Mali like settings (*long season*) as recommended by the WHO ^3^ (Fig. S2). We assume a decrease in SMC coverage by 3% between treatments (ca. 10% over a four-doses treatment regimen), therefore lying between the two extremes of observed coverage decrease^8,9^.

The *clinical trial stage* (Fig. 1b, panel 1) was simulated in the high case management setting to account for awareness of malaria symptoms. Initial deployment coverage levels for both, SMC and LAI, were set to 100%. Follow up visits, in the form of active case detection, were implemented two weeks after every administration round of SMC-SP+AQ in both trial arm. In addition to fully effective SMC-SP+AQ, we investigated a reduced length of protection by prevalent resistance against SP in the *clinical trial stage*.

In the *implementation stages* (Fig. 1b, panel 2), we analysed the impact of switching seasonal malaria prevention strategies from SMC-SP+AQ after five years of implementation to LAIs. After five years of LAI implementation, the cumulative clinical case incidence was compared to a control setting (no switching). LAIs and SMC-SP+AQ were implemented with varying coverages between 40-100 %. Intervention cohorts, defined by the coverage in the intervention age group, were specified at the beginning of a transmission season for the whole season. An exemplary illustration of LAI and SMC-SP+AQ implementation is provided in Fig. S4.

### Intervention characteristics: SMC and LAI properties

As the decay of efficacy of LAIs is not yet known, we explored a range of possible scenarios. In both the clinical and implementation analysis, the prevention of infection by LAI was modelled as pre-erythrocytic protection from infection *E(t)* over time defined by the initial protective efficacy *E*_*0*_ [%], protective efficacy half-life of decay *L* and shape parameter *k* (Fig. S3). The decay shapes of protective efficacy were chosen such that they represent multiple development possibilities: exponential-like decay (referred to as *exponential LAIs*), malaria vaccine-like decay, namely biphasic-like decay ^35,36^ (referred to as *bi-phasic LAIs*) and sigmoidal-like decay (referred to as *sigmoidal LAIs)*. The protective efficacy decay over time E(t) for exponential and bi-phasic LAIs was modelled as either Weibull-like decay:

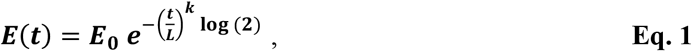

where k =1 yields *exponential LAIs* and k=0.69 yields *bi-phasic LAIs. Sigmoidal LAIs* were defined by the following Hill equation with k=8.

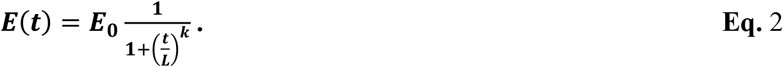

The individual protection profile over time after one administration of SMC-SP+AQ, was parameterized using published clinical trial results ^4^. The protection profile in preventing infections is not well understood ^37^, and usually the protective efficacy of SMC-SP+AQ was assessed in clinical studies in terms of population survival estimates, risk reductions or a reduction in incidence ^9,38^. To compare the impact of novel LAIs with SMC-SP+AQ, we parameterized the protective efficacy of SMC–SP+AQ to data from a clinical non-inferiority trial conducted in Burkina Faso ^4^. Our parameterization approach employs a Gaussian process regression model ^39^ to determine the model parameters that reproduce the clinical trial data via minimization of the residual sum of squares between the observed and simulated SMC-SP+AQ protective efficacy. A detailed description of the parameterization process is given in the Supplementary Information. Scenarios assuming SP resistance, the effect of resistance against the SMC component SP was implemented by decreasing the SMC half-life of protection from 32 days to 20 days. The lower protection half-life due to resistance can be modeled as an increase in the drug concentration inhibiting the parasite growth by 50% (IC_50_). Because SP has a long clearance half-life ^40^, we assume no impact of resistance on the initial efficacy.

### Health target: endpoints to assess impact of LAI

The health targets analysed in this study were based on incidence reduction by LAIs, clinical cases averted by LAIs compared to SMC, and non-inferiority with regard to clinical burden in the modelled clinical trials. Clinical cases per person in the intervention age group (*cpp*_*0*.*25-5y, int*_) over the clinical trial length (Fig. 1a) (*cpp*_*0*.*25-5y, int*_) or in the last implementation year (*cpppy*_*0*.*25-5y, int*_) (Fig. 1b) were calculated over the whole population at risk in the intervention age-group (*N*_*int,0*.*25-5y*_).

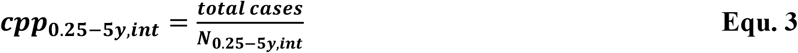

The incidence reduction percentage *inc*_*red*_ was calculated via the cumulative incidence in the year before trial implementation *cpp*_*cont*_ and the cumulative incidence during the clinical trial *cpp*_*int*_

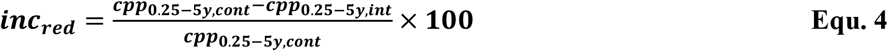

Survival analysis on the number of clinical cases per person *cpp*_*int*_ was performed to analyse the establishment of non-inferiority of LAIs to SMC–SP+AQ as described in ^41^. The impact of a LAI is assumed to be non-inferior to SMC–SP+AQ if the upper limit of the derived 95% confidence interval of the hazard ratio between SMC–SP+AQ and LAI lies below the upper limit for non-inferiority. This limit is informed by the survival estimate of SMC–SP+AQ and the desired margin of non-inferiority. We assumed a margin of non-inferiority of 5%. More information about the non-inferiority analysis is provided in the Supplementary Information.

Additionally, the intervention impact defined as clinical incidence difference between SMC-SP+AQ clinical cases per person per year *cpp*_*0*.*25-5y, SMC*_ and LAIs *cpp*_*0*.*25-5y, LAI*_ was compared using the relative difference *diff*_*cpp*_ as an indicator for the malaria burden.

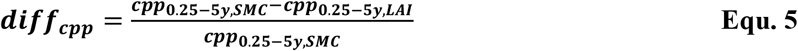

### Gaussian process emulator approach to predict intervention impact

To perform a fast and efficient search of the parameter space of LAI properties, we used a database of *OpenMalaria* simulations to train heteroskedastic GPs for each LAI efficacy decay type in each seasonal and transmission intensity setting (R-package *hetGP*, function *mleHetGP*, Version 1.1.2.) ^42^. The input parameters of the GPs in the *clinical trial stage* consisted of the tool properties including initial protective efficacy and protective efficacy half-life. In the *implementation stage*, the input parameters additionally included the respective SMC-SP+AQ and LAI deployment coverage. To define the covariance structure of the Gaussian process models, we used a Matérn kernel with smoothness parameter 5/2 accounting for the high variability in the parameter space ^42^. The trained GPs were then used to predict the number of cases per person per year and the metrics related to the non-inferiority analysis (Table S2) for any point in the parameter space. Emulator performance was ascertained by testing on a 20% holdout set after training on 80% of the data ^20^ (Table S2).

### Sensitivity analysis

We identified the main drivers of intervention impact via a global sensitivity analysis, performed using decomposition of variance via Sobol analysis on the emulator output predictions. We conducted an EIR-stratified sensitivity analysis to assess the potential change in drivers of impact over the whole transmission range. Within the pre-defined parameter space, the total effect indices quantify the interactions between individual parameter contributions to the emulator output variance ^43^. The total effect indices were normalized to obtain the relative importance of each parameter through division by their sum. The total effect indices were estimated with a Monte Carlo sampling approach using the function *soboljansen* in the R-packages *sensitivity* (version 1.16.2) with 500 000 sampled points and 1000 bootstrap replicates.

### Intervention properties and coverage optimisation

In the *clinical trial stage*, we discretized the space of the initial protective efficacy and protective efficacy half-life of LAIs within the range of plausible values (Table 1), yielding a two-dimensional grid of parameter value combinations. At each grid point, we used the GP emulators to estimate the upper limit of the confidence interval of the hazard ratio between the survival estimates in the SMC and LAI arm and the non-inferiority margin and check if non-inferiority could be established for the given combination of parameter values (cf. Non-inferiority Analysis in Supplementary Material). The contours of the resulting non-inferiority surface yielded the thresholds of the minimal initial protective efficacy and protective efficacy half-life needed to establish non-inferiority across different transmission settings (Fig. 3).

In the *implementation stage*, we additionally determined the minimum required LAI coverage and half-life at which non-inferiority to SMC-SP+AQ could be established. At each grid point, we conducted a constrained optimization, translating the non-inferiority condition into an inequality constraint by requiring the difference between the upper limit of the confidence interval and the non-inferiority margin to be positive. The optimization was conducted using an augmented Lagrange method (*gosolnp*, R-package *Rsolnp*, Version 1.16) with a minimum of 3 starting values and 200 simulations. To determine the benefits of reallocation of resources from reduced visits within a season towards increasing deployment coverage, we compared the number of cases per person per year in a simulation with SMC implemented at a given coverage to the number of cases per person per year with LAI at a coverage that was 20% higher than the optimal coverage.

## 4. Results

### Decay shape and duration of protective efficacy influence LAI impact

We assessed the obtained malaria incidence reduction in the simulated scenarios of the *clinical trial stage* at high deployment coverage (100%) and found that the decay shape of LAI protective efficacy and protection half-life play an important role in achieving a targeted incidence reduction (Fig. 2) in each simulated clinical trial scenario. Additionally, we identified the parameter space in which a certain incidence reduction cannot be achieved following LAI deployment under different setting assumptions (below each curve in Fig. 2).

**Figure 2:**
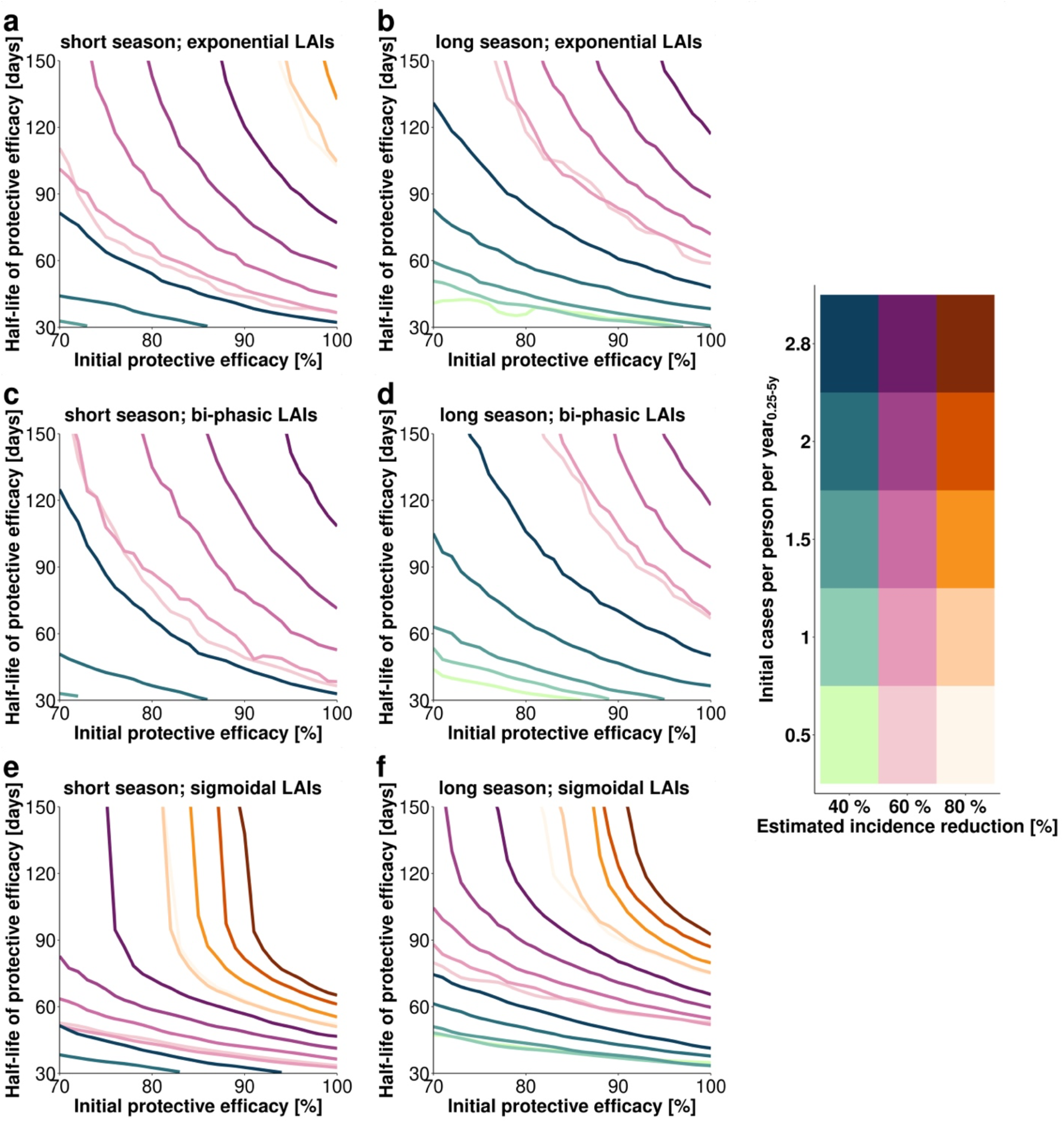
Achieving targeted malaria incidence reduction depends on the decay shape of the LAIs protective efficacy. Estimated relationships between initial protective efficacy and efficacy half-life for different incidence reduction criteria (40%, 60% and 80%, line style and color) and clinical incidence settings (increasing color intensity indicates an initial clinical incidence measured in cases per person per year in the target age group of 0.5, 1, 1.5, 2, and 2.8). Each line shows the minimum required LAI characteristics to reach the desired health goal at a 100% LAI deployment coverage at *clinical trial stage*, with all parameter combinations below a line failing to meet those requirements. The panels show the parameter space of attainable incidence reductions within the specified constrained ranges of initial protective efficacy and half-life for *exponential LAIs* (a, b), *bi-phasic LAIs* (c, d) and *sigmoidal LAIs* (e, f) in settings with a short (Senegal-like a, c, e) or long (Mali-like b, d, f) malaria season. The incidence reduction was calculated by comparing the incidence over one transmission season after application of the LAI compared to the previous transmission season. The incidence reductions were obtained by predicting the cases per person per year_0.25-5y_ via our emulator approach (See Methods and Supplementary Information) in a fine grid defined over the parameter space (increments of 0.1 h for half-life and 0.01% for initial protective efficacy) and calculating the incidence reduction by comparison to the initial clinical incidence measured in cases per person per year_0.25-5y_ in the respective transmission intensity setting.

In settings with a more extended transmission season (Fig. 2 b, d and f), a longer LAI half-life is required to reach the same predicted impact compared to the shorter season settings (Fig. 2 a, c, and e). A steeper initial decrease of initial protective efficacy (Fig. 1a and Fig. S3) led to lower estimated incidence reduction, with *bi-phasic LAIs* exhibiting the lowest predicted impact (Fig. 2 c and d), followed by *exponential LAIs* (Fig. 2 a and b). The predicted impact of *sigmoidal LAIs* was largely determined by their half-life compared to initial protective efficacy: an initial decrease in protective efficacy would require only a marginal increase of the half-life needed to reach the desired incident reduction (Fig. 2 e and f). In contrast to *exponential* (Fig. 2 a and b) and *bi-phasic LAIs* (Fig. 2 c and d), where the force of infection increased the required minimum half-life of protective efficacy, the required minimum half-life for *sigmoidal LAIs* was not significantly increased by the force of transmission (Fig. 2 e and f). A longer malaria transmission season (Fig. 2 b, d, and f) was predicted to increase the half-life requirements to reach a desired incidence reduction for all LAIs. In these longer transmission season settings, a predicted incidence reduction of over 80% was not possible for *exponential* and *bi-phasic* LAIs.

Fig. 2 shows the exemplary extraction of minimum essential efficacy properties for a LAI with a given half-life. For example, if the half-life of protective efficacy of a LAI was assumed to be 150 days, we predicted that an initial protective efficacy of 88%, 96%, and 76% was required for *exponential, bi-phasic* and *sigmoidal LAIs*, respectively (Fig. 2 a, c and e) to reach a clinical incidence reduction of 60% (short malaria transmission season, initial cases per person per year_0.25-5y_ of 2.8).

### Establishing non-inferiority of LAIs to SMC-SP+AQ in clinical trials is difficult

In the SMC-SP+AQ arm of the simulated *clinical trial stages*, we found a predicted mean achievable incidence reduction of approximatively 90 % in Mali and Senegal-like seasonal settings (Table S 3). Our non-inferiority analysis (Fig. 1b, panel 3) demonstrated that the predicted establishment of non-inferiority of *sigmoidal LAIs* to SMC-SP+AQ under the assumption of 100% initial deployment coverage could only be met with LAI efficacy over 90% in both seasonal settings and half-life over 62 days in Senegal-like (short season) and 88 days in Mali-like (long season) seasonal transmission patterns (Fig. 3). In agreement with the analysis of attainable incidence reduction (Fig. 2), the predicted establishment of non-inferiority was more feasible in settings with a shorter transmission season and lower initial malaria incidence (Fig. 3 a). For settings with a lower initial incidence (between 0.5 and 1 initial cases per person per year_0.25-5y_), the parameter space where non-inferiority could be established varied more than in higher initial incidence settings. If resistance against SP was prevalent, which we modelled as a shorter duration of protection through a decrease in protective efficacy half-life to 20 days (from 32 days), *sigmoidal LAIs* were predicted to be non-inferior in a wider range of tool property combinations (Fig. 3, c and d). Nevertheless, non-inferiority could not be established in any setting for any parameterization of *exponential* and *bi-phasic LAIs* under clinical trial coverage assumptions. We conclude that the efficacy decay profiles of LAIs play an important role for reaching the defined incidence reduction goals and establishing non-inferiority to SMC-SP+AQ.

**Figure 3:**
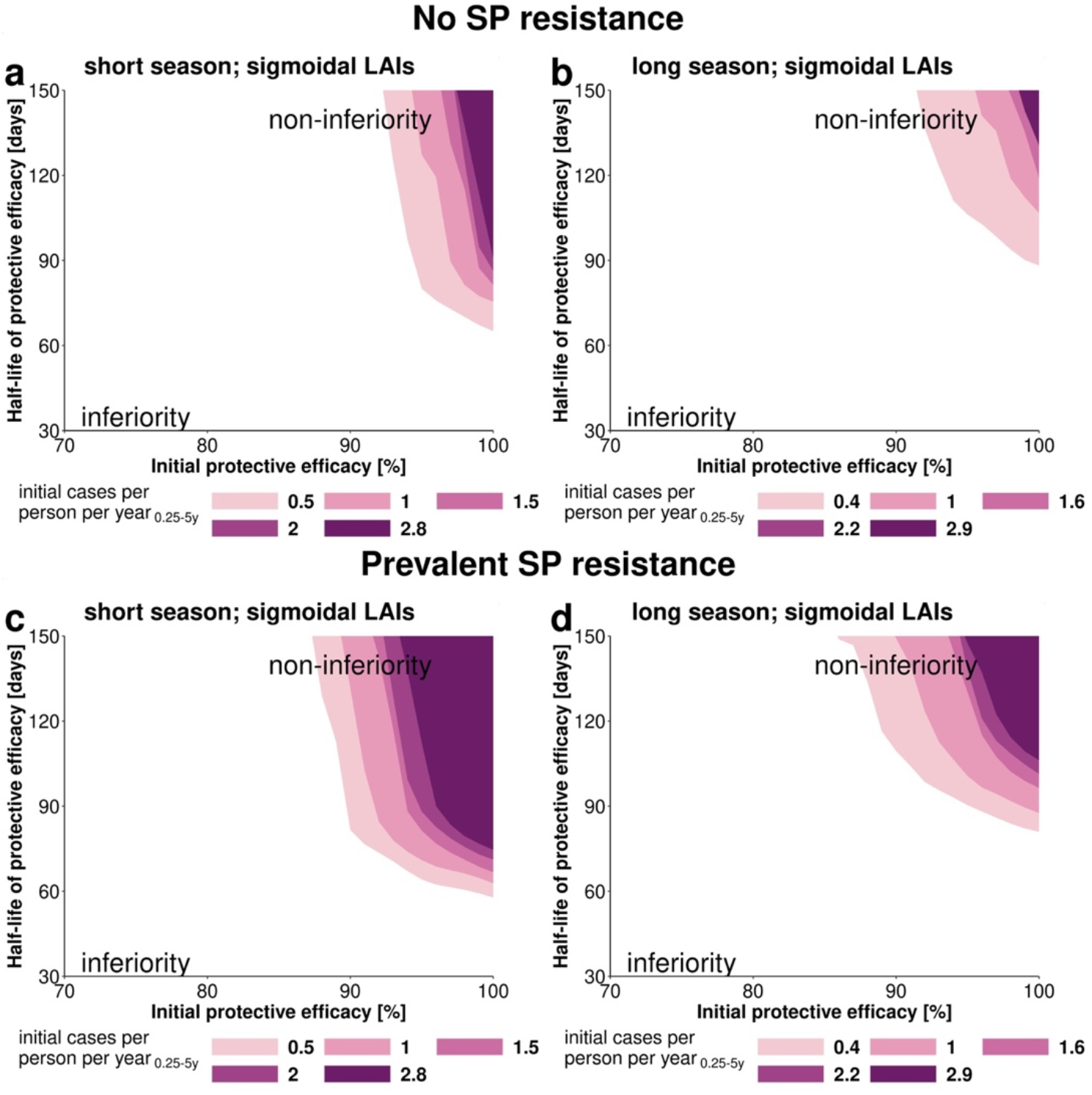
Parameter space under which *sigmoidal LAIs* achieve non-inferiority compared to SMC- SP+AQ in the *clinical trial stage*. We investigated the ability of *sigmoidal LAIs* to establish non-inferiority in clinical trials with an optimal deployment coverage (100%) in settings with a short (a, c) and long (b, d) malaria season and over varying initial malaria incidence (initial cases per person per year_0.25-5y_). (a, b) SMC–SP+AQ has an initial protective efficacy of 100% and a half-life of 32 days parameterised from previous clinical trial data^1^. The influence of prevalent SP-resistance (c, d) was analyzed by decreasing the protective efficacy half-life of SP from 32 days to 20 days (see Supplementary Information). The coloured area defines the limits of the parameter space where non-inferiority could be established through comparison of the difference hazard ratio δ between Kaplan-Meier survival estimates (see Supplementary information). The white area describes the parameter space where LAIs are inferior. *Sigmoidal LAIs* can achieve non-inferiority compared to SMC for lower durations of protection in a shorter malaria transmission season. *Exponential LAIs* and *bi-phasic LAIs* could not establish non-inferiority to SMC–SP+AQ.

### The influence of protective efficacy half-life changes over the parameter space

Moving from clinical field trials towards *implementation stages*, where LAIs are administered as a replacement for SMC-SP+AQ (Fig. 1 b, panel 2), we analyzed the influence of underlying LAI efficacy properties and deployment coverage on resulting intervention impact and non-inferiority to SMC-SP+AQ.

Our sensitivity analysis via decomposition of variance (Fig. 4) indicate that the influence of half-life of protective efficacy depends on the length of the transmission season in *implementation stages*. An increase in initial efficacy (Fig. 4 d) and deployment coverage (Fig.4 e) resulted in a linear increase in predicted impact. In contrast, the influence of the protective efficacy half-life changed over the parameter space (Fig. 4 a, b and c), with a much steeper influence for changes in lower half-life ranges. We found a change in the main source of variance of the impact of *sigmoidal LAIs* with increasing protective efficacy half-life (shown in Fig. 4 a and b for a half-life threshold of 90 days). For instance, if the LAI half-life was less than 90 days (Fig. 4a), the protective efficacy half-life and deployment coverage shared almost equal proportions of attributable variance, accounting for 38 % to 52% of variance while the initial protective efficacy contributed between 7% to 19%, depending on transmission intensity. However, for half-lives greater than 90 days (Fig. 4 b) the importance of deployment coverage increased from 54% to 85% and importance of the initial protective efficacy from 14% to 38 % (in these results we assumed LAIs of less than 70% initial protective efficacy are unlikely to be developed). In contrast, the relative importance of the protective efficacy half-life decreased to around 2% to 12%. In settings with a longer malaria transmission season (Fig. S6 a, d, g), a sharper initial decrease in clinical incidence was predicted for a larger range of protective efficacy half-life than for shorter transmission seasons (Fig. S5 a, d, g). Overall, this demonstrates the potential to explore how the impact determinants and their importance change based on efficacy duration cut-offs compared to length of transmission season or alternative deployment.

**Figure 4:**
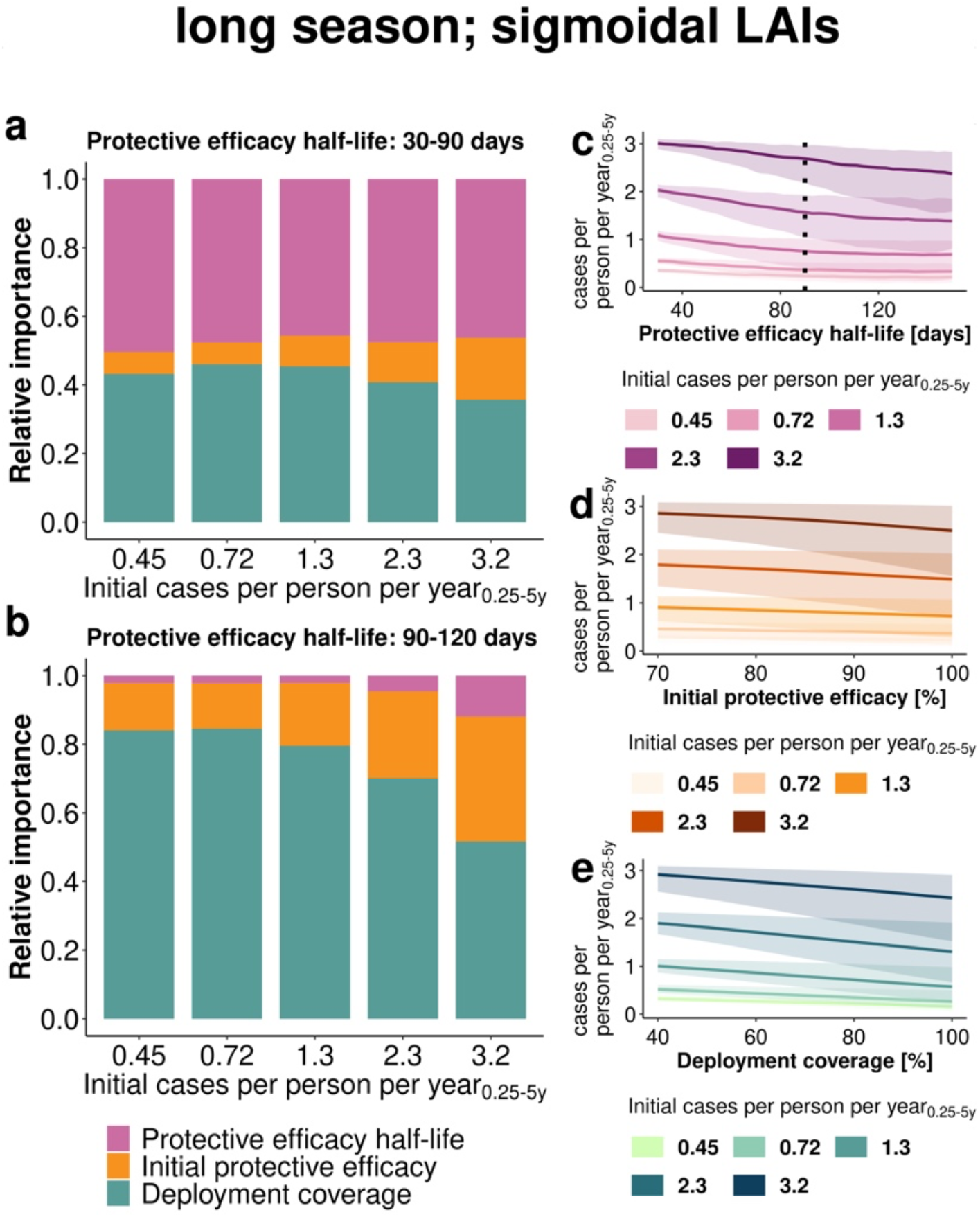
Estimated importance of LAI properties and operational factors on the level of clinical incidence reduction. Results are shown for the *implementation stage* for *sigmoidal LAIs* in a setting with low access to care and long malaria transmission season. **(a, b)** Sobol sensitivity analysis estimates the relative importance of LAI coverage, initial protective efficacy and half-life of protective efficacy to the variance of the emulator through decomposition of variance over the entire evaluated parameter space (coverage 40-100%, initial efficacy 70-100% and half-life **(a)** 30-90 days and **(b)** 90-150 days). Changes in clinical incidence measured as cases per person per year_0.25-5y_ with increasing tool properties or deployment coverage across the parameter space are shown for **(c)** half-life (30-150 days), **(d)** initial efficacy (80-100%) and **(e)** coverage (40-100 %). The lines represent the mean and the 95%-confidence bands (shaded area) capture the distribution of incidence reduction across all sampled values. The dotted line in panel c indicates the split of half-life range for sensitivity analysis in panel a and b. Increasing color intensity represents increasing initial cases per person per year_0.25-5y_. Further results for different decay shapes and length of transmission season are shown in the Supplementary Figures S4 and S5.

### Trade-offs between enhancing duration of protection and implementation coverage

These results illustrate the importance of setting-specific trade-offs between enhancing tool properties or improving implementation coverage (Table 2). For example, increasing the half-life of a *sigmoidal LAI* with an initial efficacy of 90% from 49 days to 63 days reduced the predicted required LAI coverage to establish non-inferiority to SMC-SP+AQ in implementation (60 % coverage) by 20 % (from 100 % to 80 %) in a setting with an initial clinical incidence of 1.4 cases per person per year_0.25-5y_. Furthermore, in settings with relatively high levels of initial clinical malaria incidence and corresponding high transmission intensity, namely cases per person per year_0.25-5y_>2.4 and EIR>150, a change in dynamics to establish non-inferiority was observed. In these settings, we predicted LAIs will likely fail to sufficiently protect the targeted population from clinical malaria even at very high deployment coverage (Fig. S4). Therefore, we were unable to assess the required half-life of protective efficacy of LAI for high transmission settings.

**Table 2:** Illustration of the trade-offs between LAI protective efficacy half-life, initial protective efficacy and coverage in *implementation stages*. The table displays the estimated minimum half-life of LAI protective efficacy (measured in days, estimated values specified in the colored cells, increasing colour intensity illustrates increasing requirements) required to reach non-inferiority in implementation stages of various LAI profiles compared to SMC-SP+AQ deployed at 60% coverage for each of 3 or 4 rounds in a setting with low access to health care (E_14_ = 0.1). SMC-SP-AQ protective efficacy specifications are summarized in Table 1. Results are shown for different levels of LAI coverage, decay shapes (exponential or sigmoidal), initial protective efficacy and malaria incidence prior to deployment (initial cases per person per year_0.25-5y_).

| Efficacy [%] | initial cases per person per year<sub>0.25-5y</sub> | Exponential LAIs<br>Coverage [%] |  |  |  | Sigmoidal LAIs<br>Coverage [%] |  |  |  |
| --- | --- | --- | --- | --- | --- | --- | --- | --- | --- |
|  |  | 40% | 60% | 80% | 100% | 40% | 60% | 80% | 100% |
| 70% | 0.41 | - | - | 113 | 62 | - | - | 85 | 59 |
|  | 0.71 | - | - | - | 70 | - | - | 87 | 63 |
|  | 1.4 | - | - | - | 89 | - | - | 103 | 63 |
|  | 2.4 | - | - | - | 93 | - | - | 105 | 79 |
|  | 3.3 | - | - | - | - | - | - | - | - |
| 80% | 0.41 | - | - | 109 | 54 | - | - | 71 | 52 |
|  | 0.71 | - | - | 112 | 62 | - | 148 | 74 | 55 |
|  | 1.4 | - | - | 132 | 64 | - | - | 77 | 57 |
|  | 2.4 | - | - | 119 | 75 | - | - | 84 | 62 |
|  | 3.3 | - | - | - | - | - | - | - | - |
| 90% | 0.41 | - | - | 81 | 48 | - | 102 | 61 | 50 |
|  | 0.71 | - | - | 79 | 52 | - | 86 | 64 | 52 |
|  | 1.4 | - | - | 90 | 56 | - | 103 | 63 | 49 |
|  | 2.4 | - | - | 92 | 62 | - | 97 | 72 | 56 |
|  | 3.3 | - | - | - | - | - | - | - | - |
| 100% | 0.41 | - | 112 | 62 | 46 | - | 78 | 58 | 47 |
|  | 0.71 | - | 115 | 69 | 48 | - | 78 | 58 | 49 |
|  | 1.4 | - | 131 | 71 | 49 | - | 78 | 58 | 47 |
|  | 2.4 | - | 119 | 72 | 52 | - | 80 | 61 | 47 |
|  | 3.3 | - | - | - | - | - | - | - | - |

For SMC-SP+AQ, we estimated that an additional 13 % to 29 % in incidence reduction could be achieved by increasing the coverage from 62% to 100%, dependent on initial clinical incidence before implementation (Fig. S7). However, achieving high levels of SMC coverage at implementation is challenging^5,6^, and increasing levels of coverage are associated with increasing costs.

As information on costs of LAI (costs of goods and supply chain) are not available as of now, we were unable to include detailed economic analysis in the assessment of LAIs in *implementation stages*. The main cost drivers of SMC-SP+AQ are deployment costs (remuneration of health care workers) and cost of goods ^44,45^, with deployment costs increasing non-linearly with higher coverage. Therefore, we determined the minimal LAI coverage at which non-inferiority to SMC-SP+AQ (assuming initial SMC deployment coverage of 60%) was established, stratified by initial clinical incidence before implementation (Fig 5 and Fig S8 and S9). We found that the parameter space where non-inferiority could be established shrank with increasing baseline malaria incidence (Fig. S8 and S9). With regard to seasonality, LAIs were more likely to be non-inferior in shorter malaria transmission settings in the *implementation stage* (Fig. S7 and Fig. S8). In settings with a high initial clinical incidence (EIR=150, Table S1), LAI coverage could not be optimized to be non-inferior to SMC-SP+AQ at 60% coverage because LAIs were unable to prevent malaria cases even at very high coverage levels (Fig. S4d). Overall, the optimisation of LAI coverage in settings with a limited SMC-SP+AQ coverage illustrates the potential of LAI implementation in non-optimal coverage settings.

**Figure 5:**
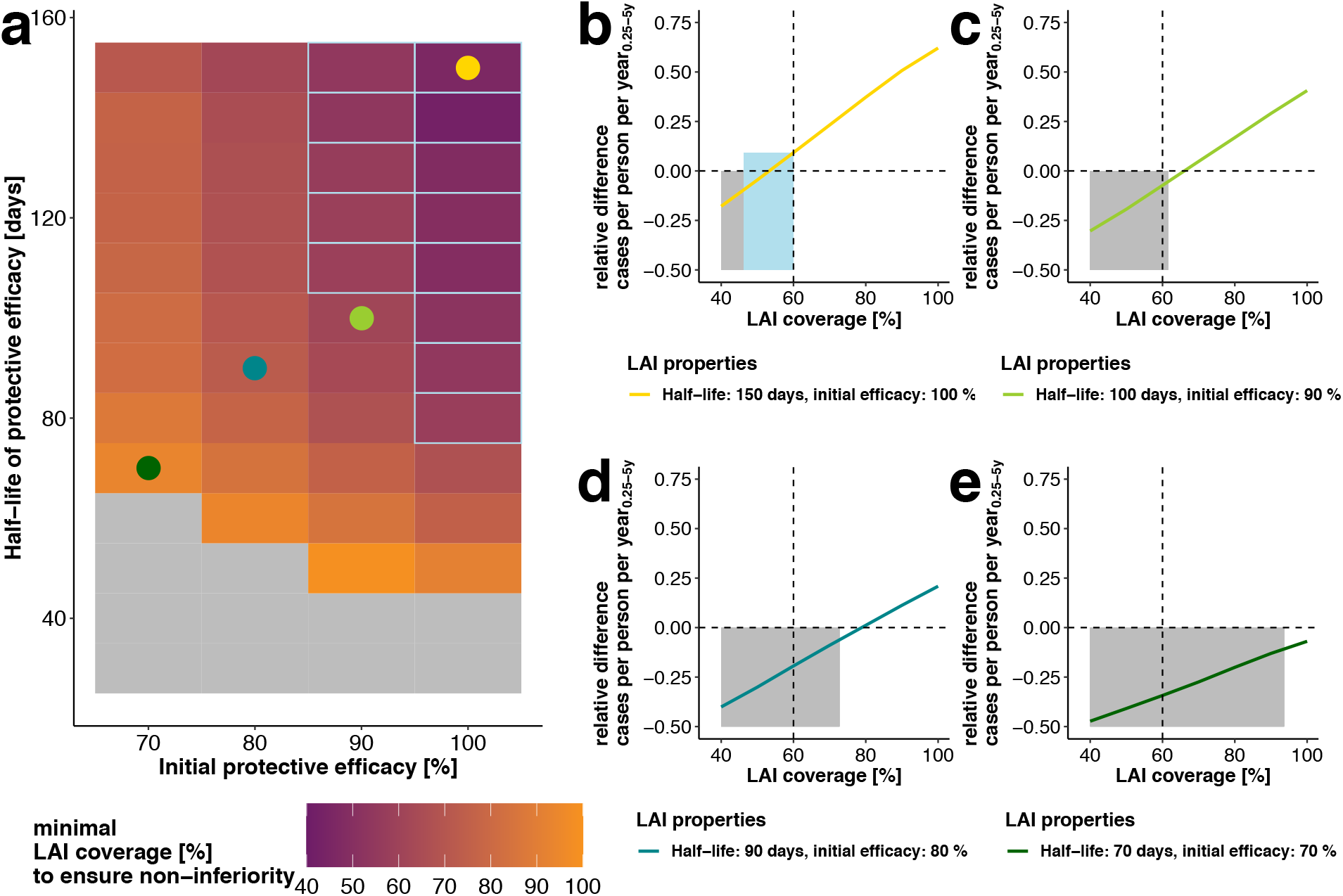
Estimated minimal LAI coverage required during *implementation stages* to achieve non-inferiority in a given setting and predicted gains in cases averted of subsequent *sigmoidal* LAI coverage increments. **(a)** Heatmap of the estimated minimal coverage (colour) of *sigmoidal LAIs* at which non-inferiority to SMC-SP+AQ (assuming a fixed SMC coverage of 60%) is achieved for different combinations of *sigmoidal LAI* efficacy and half-life. The results are displayed for intervention scenarios with an underlying disease burden of 1.4 cases per person per year_0.25-5y_, long malaria transmission season and low access to treatment (E_14_=0.1). In the grey area, non-inferiority of LAIs could not be established for any coverage. The light blue frames capture the tool characteristics where non-inferiority could be reached with a LAI coverage under the reference SMC-SP+AQ coverage of 60%. Further results for additional settings and decay shapes are provided in the supplement (Fig. S8 and S9). The coloured dots represent four illustrative LAI profiles for which the corresponding predicted relative differences in cases per person per year_0.25-5y_ (Eq. 5) are calculated in **(b-e)** five years after LAI introduction over all LAI coverages as compared to SMC-SP+AQ at 60% coverage (vertical dotted line). The predicted positive increase in relative difference in yearly clinical cases (above the dotted horizontal line) means more clinical cases are averted with LAIs than with SMC-SP+AQ. It thus illustrates the benefit of increasing *sigmoidal LAI*-coverage above the minimal required coverage to achieve non-inferiority (shown by the grey coloured area). Due to the chosen margin of non-inferiority (here 5%, see Material and Methods), LAIs are non-inferior for a slight negative relative difference in cases per person per year_0.25-5y_. In the light-blue area in (b), a LAI coverage lower than the SMC-SP+AQ coverage is sufficient to establish non-inferiority. The corresponding analysis for *exponential LAIs* can be found in Fig. S10.

While non-inferiority could only be established in a small part of the parameter space of *sigmoidal LAIs* in long seasonal transmission settings (Fig. 3b), it is possible to increase the potential area of applicability of *sigmoidal LAIs* by optimising their deployment coverage (Fig. 5a, initial cases per person per year_0.25-5y_=1.4). Deploying a sigmoidal *LAI* at 46% coverage with a half-life of 150 days and initial efficacy of 100% is sufficient to establish non-inferiority over SMC-SP+AQ at 60% coverage (Fig. 5 b). In contrast, *sigmoidal LAIs* with a half-life of 70 days and initial efficacy of 70 % require a deployment coverage of 95% in order to be non-inferior (Fig. 5 e). For *sigmoidal LAIs* we found that increasing the deployment coverage over the estimated minimum required coverage to establish non-inferiority results in potential gains in terms of clinical incidence reduction compared to SMC-SP+AQ (Fig. 5 b to e).

Additionally, we found that even though non-inferiority of *exponential LAIs* to SMC-SP+AQ could not be established in *clinical trial stages*, coverage optimisation in *implementation stages* reveals their applicability. Deploying an *exponential LAI* at 52% coverage with a half-life of 150 days and initial efficacy of 100% was sufficient to establish non-inferiority over SMC-SP+AQ at 60% coverage (Fig. S10b) and a half-life of 100 days and initial protective efficacy of 90% requires a deployment of 78% to establish non-inferiority (Fig. S10c). *Exponential LAIs* with a half-life of 70 days and initial efficacy of 70 % were always inferior to SMC-SP+AQ at a coverage of 60% (Fig. S10e).

## 5. Discussion

The effective prevention of clinical malaria in children is crucial to prevent malaria mortality and reduce the overall global malaria burden ^1^. Through modelling and simulation, we explored a broad range of LAI characteristics in multiple settings for clinical testing and deployment. This allowed us to understand the likely influence of LAI efficacy properties and operational factors on clinical incidence reduction in children under five years of age when LAI is deployed as a seasonal malaria prevention tool. We found that if the protective efficacy of a new LAI decays immediately after injection, for example an exponential or bi-phasic-like decay, then the LAI is unlikely to achieve non-inferiority over SMC-SP-AQ in current SMC settings in a clinical trial. This exploration assumed non-inferiority criterion is required for testing, and we only explored LAI half-life of protection in the range of 30 to 150 days. In contrast, when the protective efficacy of LAIs is long-lasting and decays only after some time (i.e. a sigmoidal decay), there is a stronger chance of achieving non-inferiority when the duration of protection is close to the transmission length. Beyond clinical stages, by assessing implementation factors versus LAI properties, we conclude that focusing on enhancing the duration of protective efficacy (half-life) is more likely to result in successful LAIs in a larger range of incidence and transmission settings. If the half-life of protective efficacy of a LAI approaches the length of transmission season, and initial efficacy is sufficiently high (depending on the transmission intensity (Fig. 2)), then the development of new LAI should prioritize optimizing operational delivery factors to ensure reasonable coverage to be as good as or better in averting clinical cases than current SMC-SP+AQ implementation.

### The estimated impact of tool properties

In general, the duration of the half-life and its shape of decay are the most relevant tool properties for incidence reduction. This means that the development of new LAIs should focus on understanding the likely decay and half-life of protection (this is not the pharmacokinetic properties of a small molecule or the antibody longevity of a mAb). Over the analyzed transmission settings and parameter space, the estimated minimum essential (80%) and ideal (95%) incidence reduction targets for LAIs as defined by Macintyre et al. (2018)^15^ are hard to achieve in clinical testing. A desired incidence reduction of >80% in clinical testing could only be achieved for LAIs with a sigmoidal decay shape and half-life over 60 days (Senegal, short season) and 80 days (Mali, long season). Our findings indicate that key to identifying and refining candidates for development of new LAIs is investigating the decay shape of efficacy as early as possible and providing a sufficiently long protection half-life. Before conducting large scale clinical trials, it will therefore be important to ensure the adequate means to establish the decay and duration of protection and the ability to extrapolate to paediatric indications.

While the pharmacokinetic profile of potential LAIs can be evaluated in pharmacokinetic studies, their protective efficacy profile and decay shape are harder to derive. Currently, murine ^46^ or human challenge ^47,48^ studies are being used to investigate these parameters. In these studies, subjects receive treatment before inoculation with sporozoites through the bites of infectious mosquitoes or direct venous injection ^47,48^. However, parasite growth in the liver, and therefore protective efficacy, cannot be directly quantified. Instead, the time and number of parasites entering the blood stream is used as a crude proxy for protective efficacy. The approach presented here can aid to translate survival estimates from murine and human studies by estimating the protective efficacy decay curves and then plugging them into the population modelling approach with *OpenMalaria*. Hence, our approach can provide first insights into the potential public health impact of new LAIs.

In summary, our findings suggest that when defining key efficacy characteristics in TPPs for LAIs there are two important aspects to consider: i) evaluating the feasibility of currently or newly expect specified LAI efficacy and duration requirements ^15^ by estimating the likely public health impact of new LAIs with e.g. modeling and simulation approaches, and by ii) translating population efficacy on clinical cases into individual protection efficacy and vice versa via adequate summary measures of protective efficacy in animal or human studies.

### Clinical non-inferiority trials

Our non-inferiority analysis (Fig. 3) highlights that the establishment of non-inferiority in *clinical trial stages* is challenging due to not only the high protective ability of SMC-SP+AQ but also clinical trial designs. This motivates an important discussion on the clinical development of LAIs under the use-case of SMC replacement: the necessity and extent of clinical trial testing or non-inferiority criteria should be assessed. We estimated SMC-SP+AQ in a clinical study results in incidence reductions of 71% to 90% in children under five, aligning to a efficacy of 86% (95% CI 78–91%) in a clinical trial in Senegal ^49^. Monthly SMC-SP+AQ administration together with the estimated protective efficacy half-life of 32 days offers a high degree of protection and the blood-stage clearing effect of AQ eliminates remnant malaria infections, making optimally employed SMC a very powerful tool to prevent clinical malaria. This means it is difficult for most LAIs to achieve non-inferiority in a clinical trial with limited SP resistance when the LAI is only deployed once per season in combination with a blood stage clearing drug. Thus, at first glance, our non-inferiority results may offer a distorted picture of the ability of LAIs to compete with current SMC, as clinical non-inferiority trials do not reflect the reality of SMC implementation. Operational constraints result in reduced coverage over the number of rounds ^5,6^ and adherence ^10^, thus hindering SMC with SP+AQ from reaching its full potential. AS SMC-SP+AQ effectiveness reduces with operational challenges, this presents a niche for LAIs. In 2019 only 62% of eligible children received SMC-SP+AQ ^1^, meaning that additional incidence reduction could be achieved by increasing coverage at each round.

### Tool and coverage optimization in implementation stages

Replacing SMC-SP+AQ with LAIs will likely prove beneficial in reducing deployment costs due to fewer deployment rounds. In the absence of information on costs of LAIs (costs of goods and supply chain), we are unable to adequately assess economic considerations and thus our results assume that coverage is the main driver of implementation cost. If LAIs are assumed to have a longer clearance half-life and therefore higher protective efficacy for longer than SP+AQ, resources are freed up through decreased deployment rounds within a season. These resources could be reallocated to increase the overall coverage in a single round of LAI in the target population, including populations in remote places. Additionally, the overall adherence to the blood-stage clearing co-administration of antimalarials could be increased, thereby reducing the probability of emergence of resistance. However, if transmission intensity is very high, we found that the optimization of protective efficacy half-life and deployment coverage is insufficient to adequately protect the targeted population. Instead, it might be necessary to expand the deployment of LAIs to multiple administration rounds within a transmission season.

The optimization of deployment coverage of LAIs to reach non-inferiority to SMC-SP+AQ where optimal coverage cannot be met exposes an additional use case for LAIs. If external circumstances, such as the current COVID-19 pandemic, prevent the regular implementation of SMC and bed-nets campaigns, millions of children will experience an increased risk of malaria ^50^, LAIs may alleviate this burden. Our analysis can aid the identification of minimal LAI coverages necessary to achieve given population impacts and prevent a resurgence in malaria cases.

### Framework and areas of application

Our modelling and simulation analysis provides important insights into the likely impact of new malaria tools that are currently under clinical development. Our modelling framework divides the unknown parameter space of realistic properties of new tools into setting-specific attainable incidence reduction by translating the decay of individual protection against infection of a new tool into estimates of population impact on clinical incidence. While in clinical trials often only one (high) deployment coverage and a limited number of trial-arms can be investigated, a simulation-based approach can explore the trade-off between operational constraints and tool properties to narrow down beneficial implementation settings and use cases without the need for expensive field studies. Not only does this approach offer the opportunity to assess the potential population impact of new tools currently under development, but it also provides a methodology to assess the potential clinical trial outcomes. It assists the evaluation of clinical trial scenarios that might be considered over several different malaria transmission and health-system settings to supporting thinking on appropriate population impact endpoints that are suitable to inform decision making. This is particularly true for existing interventions with high efficacy for which the establishment of non-inferiority in clinical non-inferiority trials is problematic, due to the required large sample sizes ^51^. Here, our approach offers first insights into the outcomes of such trials and the additional possibility to develop clinical trial analysis tools.

Furthermore, beyond the current scope of our study, as more information on likely costs of LAIs become available and further certainty in implementation and cold-chain needed, this work can serve as a basis for cost-effectiveness or economic analysis.

As with all modelling studies there are limitations to our analysis. In this study, despite exploring a large range of characteristics on tool properties, deployment, and transmission settings, our results are constrained by the investigated parameter-space. First, we only investigated the impact of one administration round of LAIs with an antimalarial treatment and assumed that the time-point of administration would coincide with the first SMC application round. However, depending on the LAI profile, the time of deployment may need to be optimized. Second, the implementation of protective efficacy of LAIs is solely assumption-driven, as clinical data is not yet available. With additional information on likely achievable protective efficacy half-life, initial protective efficacy and efficacy decay shapes, the preliminary LAI profiles can be further defined and reevaluated as LAIs are developed. The re-definition of plausible parameter ranges will also impact the results of the sensitivity analysis and trade-offs, and potentially shift the recommended focus of development efforts. Additionally, target mediated drug disposition might change the pharmacokinetic profile of mAbs dependent on the transmission intensity and parasite growth within the human host and therefore also its efficacy profile ^52^. Third, the focus of this analysis was the investigation of the effect of the anti-infective LAIs. Our results are subject to change if LAIs are co-administered with different blood-stage clearing drugs (different efficacies and/or potential properties e.g. transmission blocking) or are deployed with other interventions such as insecticide-treated bed-nets. And lastly, we explored SMC or LAI replacement in only children under 5 years of age and in settings similar to where SMC is currently deployed such as Mali and Senegal. Further analysis could be undertaken to assess LAI as seasonal prevention in children under 10 years of age, however we expect conclusions to be similar in regards tool properties and coverage requirements. Our results also only hold for assessing LAI as replacing SMC; we did not explore use cases of deploying LAI in perennial or other settings in which SMC is not yet deployed. Alternative clinical metrics would need to be explored as LAI in these use-cases are not a replacement tools, rather new tools and non-inferiority trials are not relevant. Although this study focuses on the use of LAIs in seasonal malaria transmission settings, our findings regarding the importance of protective efficacy half-life do provide first insights for potential use of a LAIs in perennial malaria transmission settings. The protective efficacy half-life of a LAI will most likely dictate the number of applications to children within one year to ensure effective clinical case reduction.

### Current stage of development of mABs and duration of protection

Potential candidate LAIs include mABs and small-molecule drugs, and to date most known mAbs for use in malaria (largely by-products from research into whole sporozoite vaccines ^53^) have been shown to prevent blood-stage infection in *in vitro* and/or *in vivo* murine malaria infection experiments ^18,19^. As the natural clearance half-life of antibodies ranges between 2 and 21 days ^54^, strategies to increase the half-life of mAbs have been introduced via modifications to the tail (Fc) region of antibodies that interacts with the receptors on the surface of cells. Fc-modified mAbs have exhibited extended half-lifes ranging from, 100 days ^55^ and 80 to 112 days ^56^ in healthy human adults. Our results suggest that these extended half-lifes, if functional malaria protection is maintained, are crucial to establish non-inferiority to standard SMC. A second stream of LAI development is focused around small-molecule drugs such as atovaquone ^16^ and P218 ^17^. Although these compounds show promising liver-stage activity, the estimated clearance half-life of enhanced formulation atovaquone of 32 days in humans ^16^ and 8.9-19.6 hours of P218 in first-in-human trials ^57^ are again likely insufficient in the use cases explored in our study and emphasize the need for longer-lasting formulations to be useful as SMC replacements when deployed only once per season. Further use may be possible for multiple applications within a season.

## Conclusion

Here, we provided the first quantitative evaluation of the TPPs of future LAIs for malaria as a seasonal prevention tool in children. Simulation analysis of LAIs in real-life implementation settings revealed that the ability of LAIs to prevent clinical cases in children is strongly dependent on the length of the malaria transmission season and transmission intensity. We also found it is important to focus on improving the protective efficacy duration (half-life) of LAIs in development, as the speed of protective efficacy decay is a key driver of overall impact or the chance to meet non-inferiority criteria compared to SMC-SP+AQ. However, if a reasonable duration is possible (longer half-life and sigmoidal decay that supports protection close to the length of transmission season) then development should focus on increasing deployment coverage to optimizing the LAIs chance of higher impact. This provides evidence for the potential trade-offs between tool properties and operational constraints as LAIs are developed and deployed. In general, our findings support the need for a thorough and combined investigation of tool properties and use cases in the future development of LAIs. This combined effort includes earlier modelling alongside clinical studies to provide evidence of translation of impact at population levels before late stage clinical studies and optimize the success of new malaria tools. Our research here provides an initial foundation to support dialogue between stakeholders, scientists, and clinicians at each clinical development stage of novel anti-infective LAI’s to reduce clinical malaria incidence. LAIs have the potential to be a game changer in protecting vulnerable populations from malaria. Our analysis serves as a stepping stone for the refinement of TPPs for LAIs, thereby assisting the target-oriented use-case of development and implementation of new LAIs.

## Data Availability

Simulated results are available, no clinical or preclinical data was used in this study

## List of abbreviations

SMC: Seasonal malaria chemoprevention
LAI: Long acting injectable
TPP: Target product profile
mAB: Monoclonal antibody
SP+AQ: Sulfadoxine-Pyrimethamine+Amodiaquine
GP: Gaussian process

## 6. Declarations

### Ethics approval and consent to participate

Not applicable

### Consent for publication

Not applicable

### Competing interests

J.J.M. is employed by the Medicines for Malaria Venture. All other authors declare no competing interests.

### Funding

The work was funded by the Swiss National Science Foundation through SNSF Professorship of MAP (PP00P3_170702). The funders had no role in study design, data collection and analysis, decision to publish, or preparation of the manuscript.

### Authors’ contributions

LB, MAP, and MG designed the simulation experiments and the computational framework. LB carried out the simulations, analysis and visualisation. LB, MG, TR, JJM, and MAP contributed to the interpretation of the results. LB wrote the manuscript with input from ML, TR, JJM, and MAP. MAP and LB conceived the study and were in charge of overall direction and planning. MAP supervised the project. All authors read and approved the final manuscript.

## Acknowledgments

We acknowledge and thank our colleagues in the Swiss TPH Disease Modeling unit for their valuable insights and feedback. Calculations were performed at sciCORE (http://scicore.unibas.ch/) scientific computing center at University of Basel.

## Supplementary material for

## 1. General OpenMalaria specification and intervention set-up

**Figure S 1:**
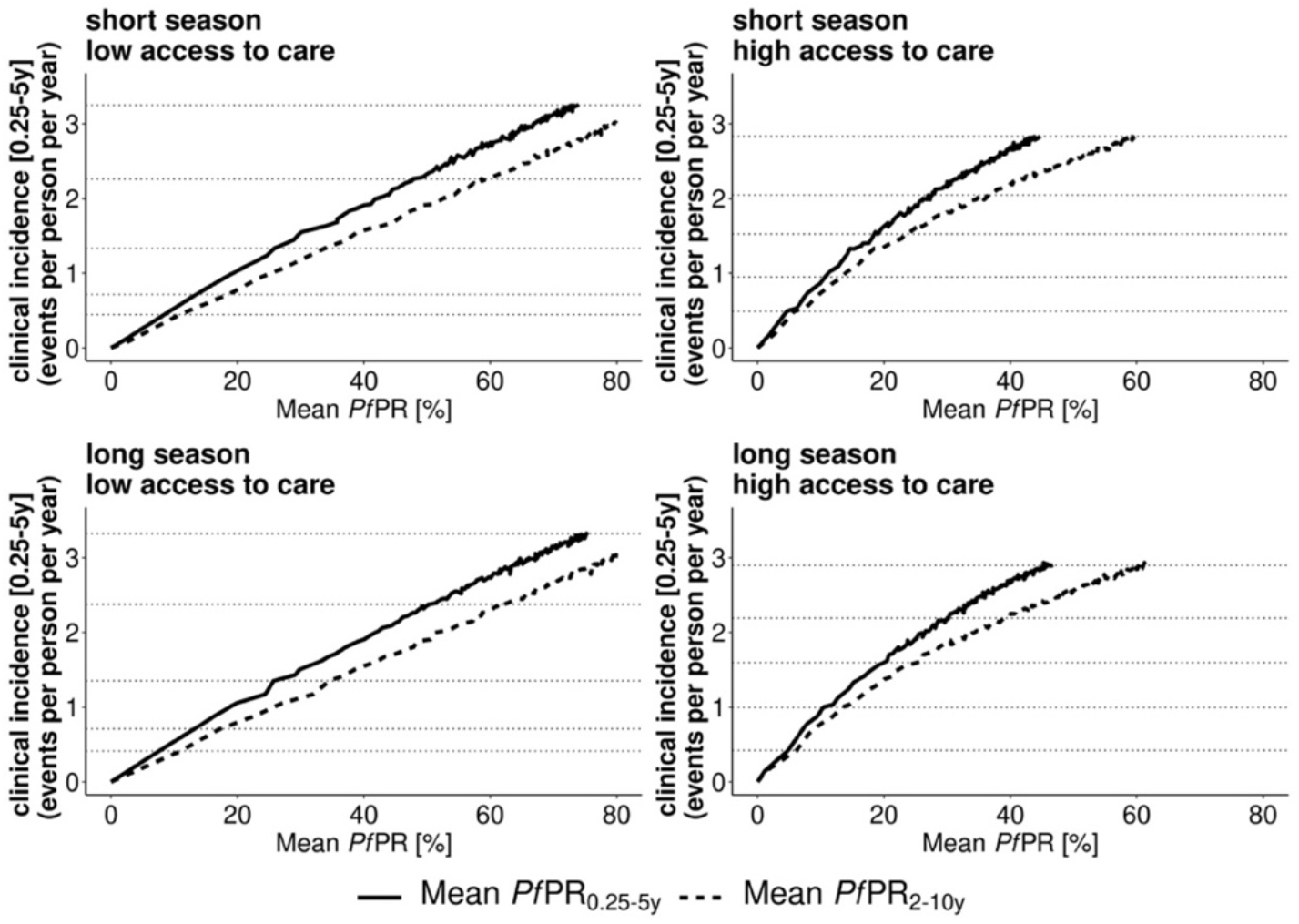
Prevalence - incidence relationship in the simulated Open Malaria settings. The relationship between prevalence and incidence is displayed in the two simulated seasonal settings (Senegal/short season and Mali/long season) and health system access settings (low and high access) (Table 1) in absence of any interventions. The clinical incidence defined as the events per person per year in the target age-group (0.25-5 years of age) is shown for the corresponding mean prevalence over one year in the intervention age group (*Pf*PR_0.25-5y_) and in children between 2-10 years (*Pf*PR_2-10y_). The dotted horizontal lines mark the incidence settings, simulated for all downstream analyses and can be found in Table S1.

**Table S 1:** Simulated prevalence – incidence settings. The force of transmission was defined by the entomological inoculation rate (EIR: mean number of infectious bites per person per year (pppy)). Across different transmission settings defined by health system access, EIR levels and transmission seasonality, corresponding simulated malaria prevalence (PfPR) and clinical incidence per person per year in the intervention age group is displayed for different age groups: 2 to 10 year olds (PfPR_2-10y_) and 0.25 to 5 years old (PfPR_0.25-5y_ and cases per person per year_0.25-5y_).

| HS Access | EIR [infectious bites pppy] | Short transmission season |  |  | Long transmission season |  |  |
| --- | --- | --- | --- | --- | --- | --- | --- |
|  |  | PfPR<sub>2-10y</sub> [%] | PfPR<sub>0.25-5y</sub> [%] | Cases per person per year<sub>0.25-5y</sub> | PfPR<sub>2-10y</sub> [%] | PfPR<sub>0.25-5y</sub> [%] | Cases per person per year<sub>0.25-5y</sub> |
| high | 5 | 6 | 5 | 0.49 | 6 | 5 | 0.42 |
|  | 9 | 13 | 11 | 0.95 | 13 | 10 | 1 |
|  | 20 | 23 | 19 | 1.52 | 25 | 20 | 1.6 |
|  | 47 | 37 | 28 | 2.04 | 39 | 30 | 2.2 |
|  | 150 | 60 | 45 | 2.83 | 62 | 46 | 2.9 |
| low | 3 | 11 | 8 | 0.45 | 11 | 8 | 0.41 |
|  | 4 | 19 | 13 | 0.72 | 17 | 13 | 0.71 |
|  | 8 | 34 | 26 | 1.3 | 35 | 26 | 1.4 |
|  | 28 | 59 | 48 | 2.3 | 62 | 50 | 2.4 |
|  | 150 | 86 | 73 | 3.2 | 87 | 75 | 3.3 |

**Figure S 2:**
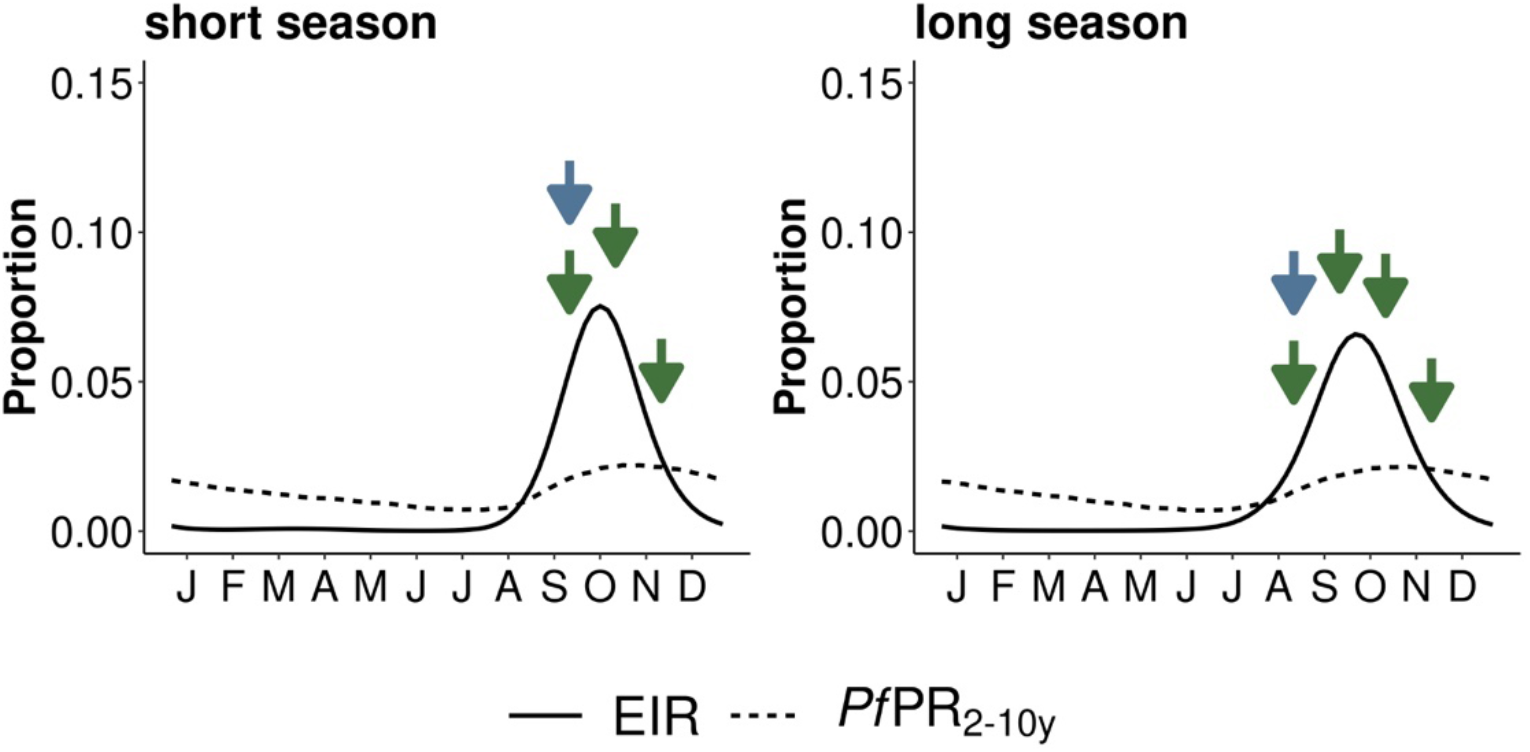
Modelled malaria transmission pattern and simulated prevalence defining seasonality settings. The seasonality in malaria transmission is defined through the proportional EIR (solid line) in relation to the yearly EIR (input into the simulator). Here shown for a yearly EIR of 100 infectious bites per person per year. The dotted line represents the resulting scaled yearly prevalence profile. The arrows illustrate the administration of the preventative interventions of SMC-SP+AQ (green) and LAI (blue) in the two seasonal settings. Following WHO recommendations^**1**^, SMC-SP+AQ was implemented monthly with the first dose administered before the peak and the second dose with the peak in malaria transmission.

**Figure S 3:**
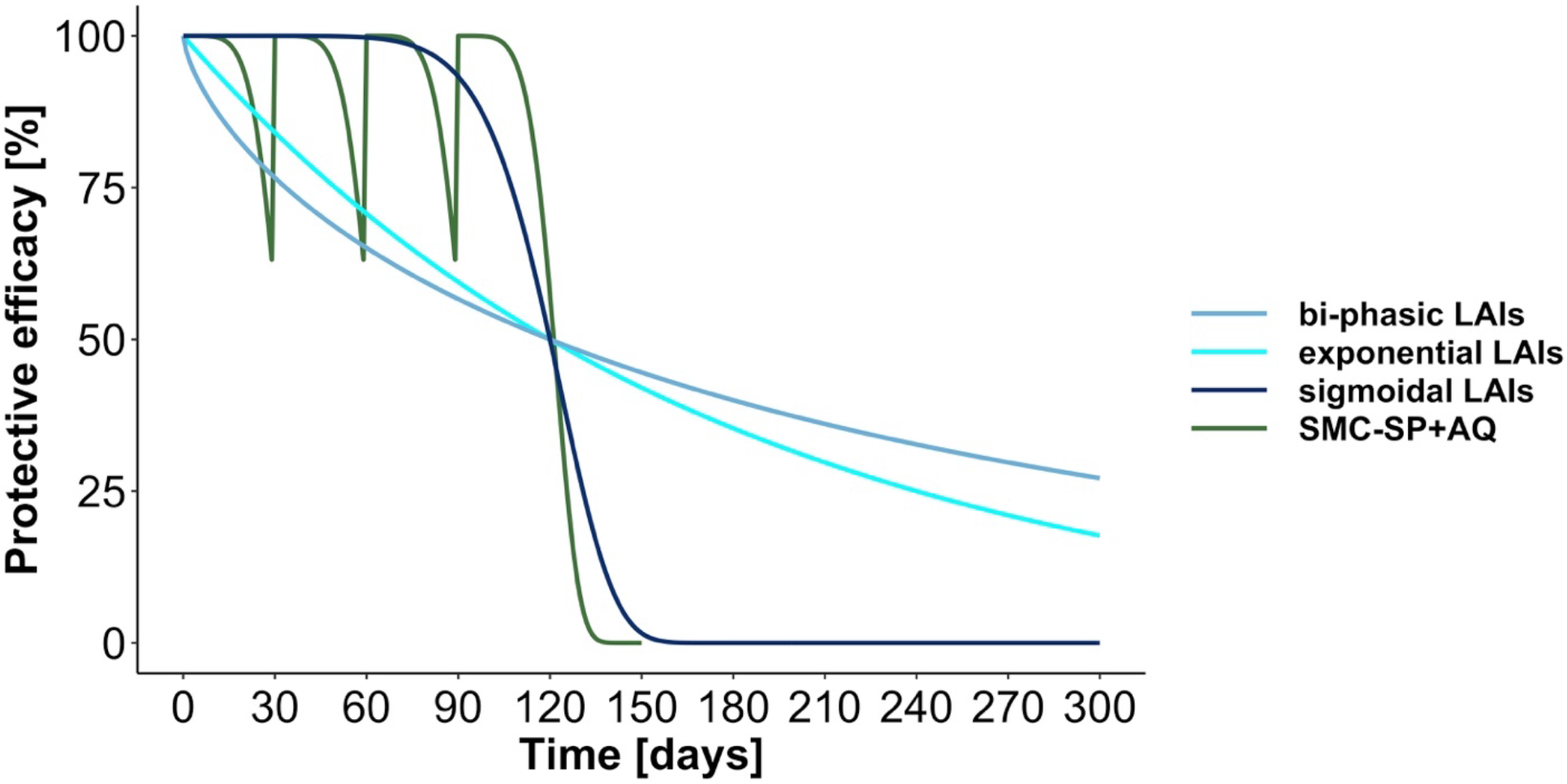
Simulated protective efficacy decay shapes of long acting injectables (LAI) and seasonal malaria chemoprevention (SMC) over one transmission season. The LAI decay shapes were chosen such that they represent the three possible development streams of mAb (sigmoid decay), drug based (exponential decay), or vaccine like (bi-phasic) LAIs (blue solid line). Here, we illustrated a LAI protective efficacy half-life of 120 days and initial protective efficacy of 100 %. SMC – SP+AQ was parameterised as specified below and administered up to four times over the transmission season (green solid line). The parameterisation of the interventions is further specified in Table 1.

**Figure S 4:**
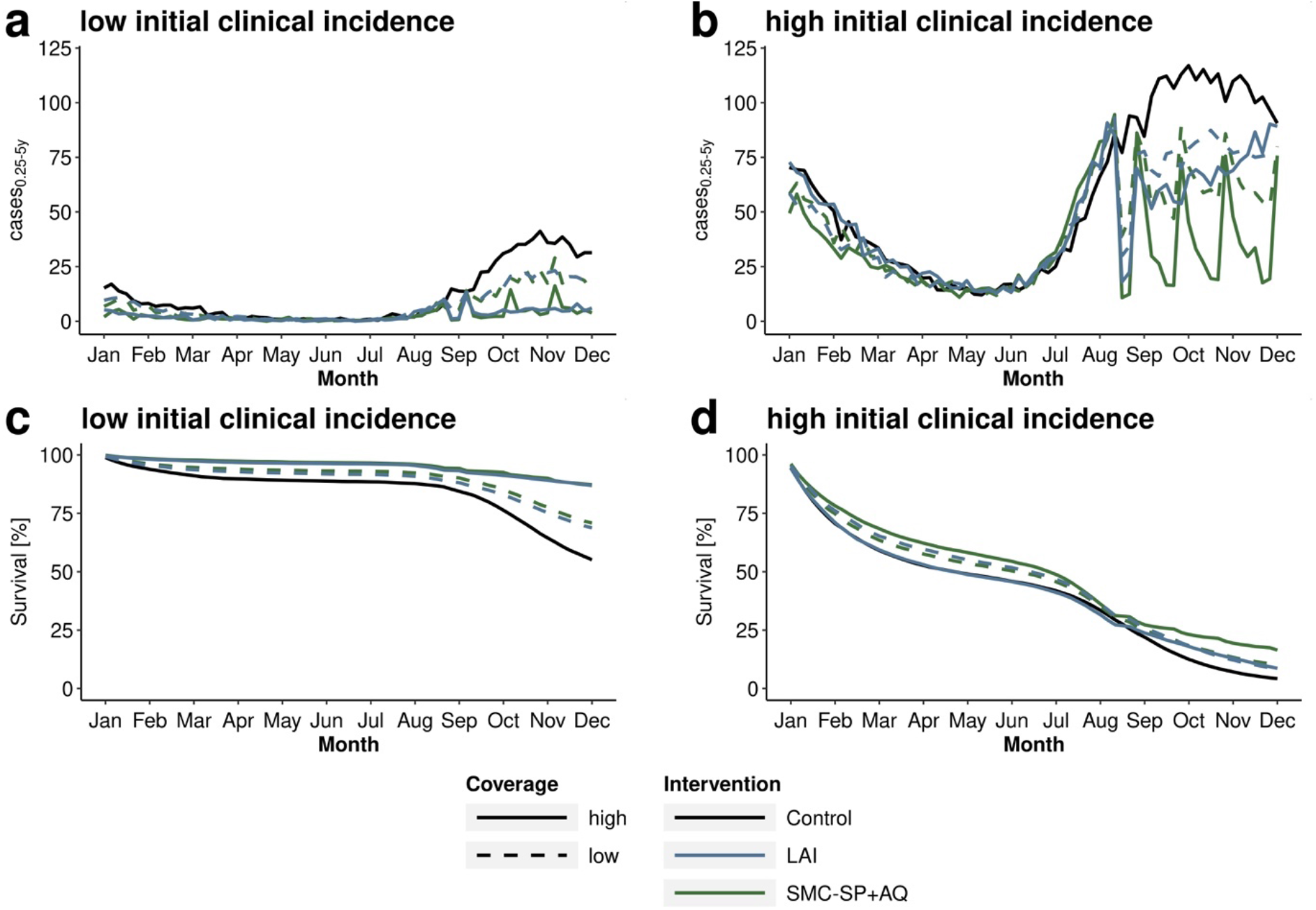
Exemplary illustration of incidence and survival estimates of *sigmoidal LAIs* and SMC- SP+AQ in *implementation stages* over one implementation year. Plots are shown in **(a, c)** low initial clinical incidence settings (initial cases per person per year_0.25-5y_ = 0.71) and **(b, d)** high initial clinical incidence settings (initial cases per person per year_0.25-5y_ = 3.3) in long malaria transmission settings with low access to health care (E_14_=0.1). The *sigmoidal LAIs* exhibit an initial protective efficacy over 95 % and protective efficacy half-life of 112 days. SMC-SP+AQ and LAIs were implemented at high deployment coverage (>90%, solid lines) and low deployment coverage (<50%, dashed lines). In low initial incidence settings **(a, c)** SMC-SP+AQ (green) is administered four times per season (Aug-Nov) with clinical incidence notably decreasing after each administration **(a, b)** compared to control settings with no implemented interventions (black). In contrast, the *sigmoidal LAIs* are only administered once at the beginning of the transmission season. With progressing season, the protective efficacy decays, resulting in an increase of clinical cases **(a, b)** and decrease in survival estimates **(c, d)** after LAI administration. In low clinical incidence settings, *sigmoidal LAIs* and SMC-SP+AQ are comparably effective at both coverage levels **(c)**. In contrast, the survival estimates in in high initial clinical incidence settings **(d)** reveal that *sigmoidal LAIs* at both coverage levels and SMC-SP+AQ at low coverage levels are all equally unable to prevent malaria cases. Shown here are the mean predictions over 5 stochastic *OpenMalaria* simulations.

## 2. Non-inferiority analysis

Survival analysis was performed using a Kaplan-Meier approach as specified in ^2^, with the K-M estimate *Ŝ*(*t*) derived via number of new clinical cases c_i_ at each time step t_i_

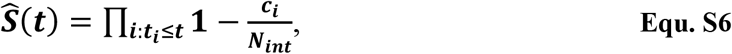

with the standard error 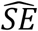 calculated using the Greenwood formula

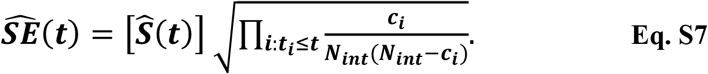

Non-inferiority analysis was conducted based on the survival statistics as described in ^2^. The survival estimate (hazard ratio) for the standard of care SMC *Ŝ*_*SMC*_ (t) considering the desired margin of non-inferiority Δ defines the upper limit for non-inferiority *γ*:

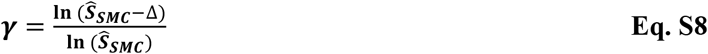

The difference δ between the survival estimates of the standard of care SMC *Ŝ*_*SMC*_ (t) and new treatment (LAI) *Ŝ*_*LAI*_, is calculated on the log-log-scale and is equivalent to the logarithm of the ratio of cumulative hazards in the two groups

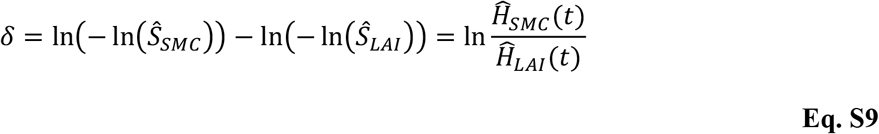

and its variance is calculated as follows:

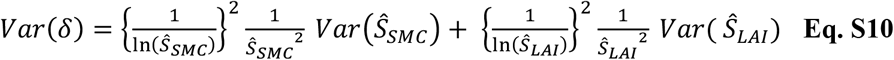

The confidence interval of the hazard ratio *δ* is given by *exp*^*δ* [95% *CI*: *δ*±1.96×*se*(*δ*)]^. Non-inferiority is established if the upper limit of the derived 95% confidence interval, CI_high_, of the hazard ratios *δ* between SMC and LAI lies below the upper limit for non-inferiority *γ*.

## 3. Emulator performance

**Table S 2:** Emulator performance for the investigated outcomes. The emulator performance was assessed by calculating the Pearson correlation coefficient (PCC) and the mean absolute error (MAE) between true and predicted values on a 20% holdout set for each investigated setting.

| Outcome | PCC<br>(median [min, max]) | MAE<br>(median [min, max]) |
| --- | --- | --- |
| upper limit for non-inferiority $\gamma$ | 0.9924 [0.8072, 0.9972] | 0.0015 [0.0002, 0.0280] |
| upper limit of the hazard ratio | 0.9945 [0.9472, 0.9978] | 0.0229 [0.0094, 0.0791] |
| Clinical incidence SMC-SP+AQ,<br>(cases per person per year<sub>0.25-5y, SMC</sub>) | 0.9964 [0.9787, 0.9986] | 0.0142 [0.0060, 0.0428] |
| Clinical incidence LAIs,<br>(cases per person per year<sub>0.25-5y, LAI</sub>) | 0.9947 [0.9562, 0.9985] | 0.0136 [0.0055, 0.0488] |

## 4. Additional analysis results in the clinical trial setting

**Table S 3:**
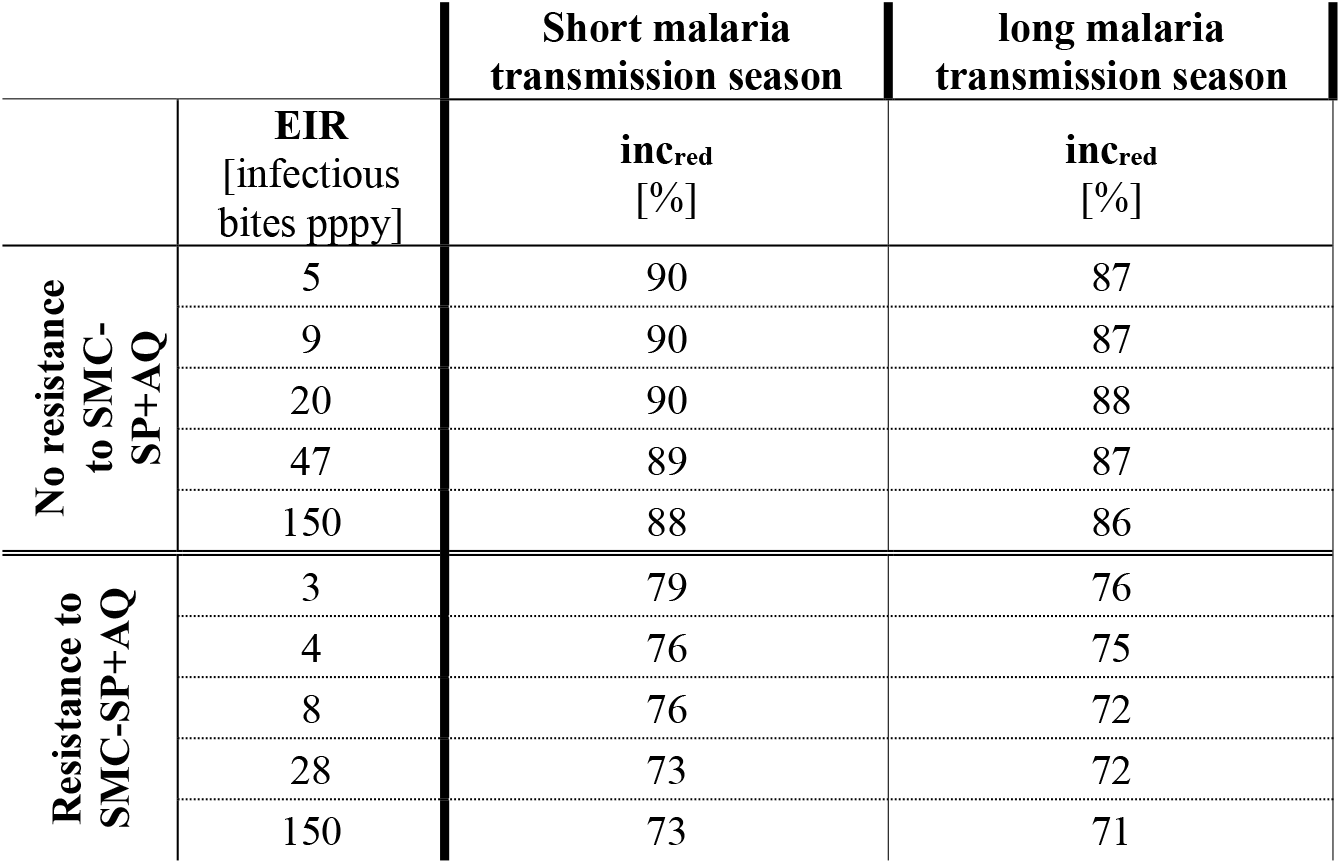
Incidence reduction achieved through implementation of SMC-SP+AQ in a clinical trial setting. The incidence reduction was calculated in the clinical trial setting over three months in the short season setting and four months in the long season setting. Resistance to SMC-SP+AQ is implemented as a reduction in protective efficacy half-life from 32 days to 20 days.

## 5. Additional analysis results in the implementation setting

**Figure S 5:**
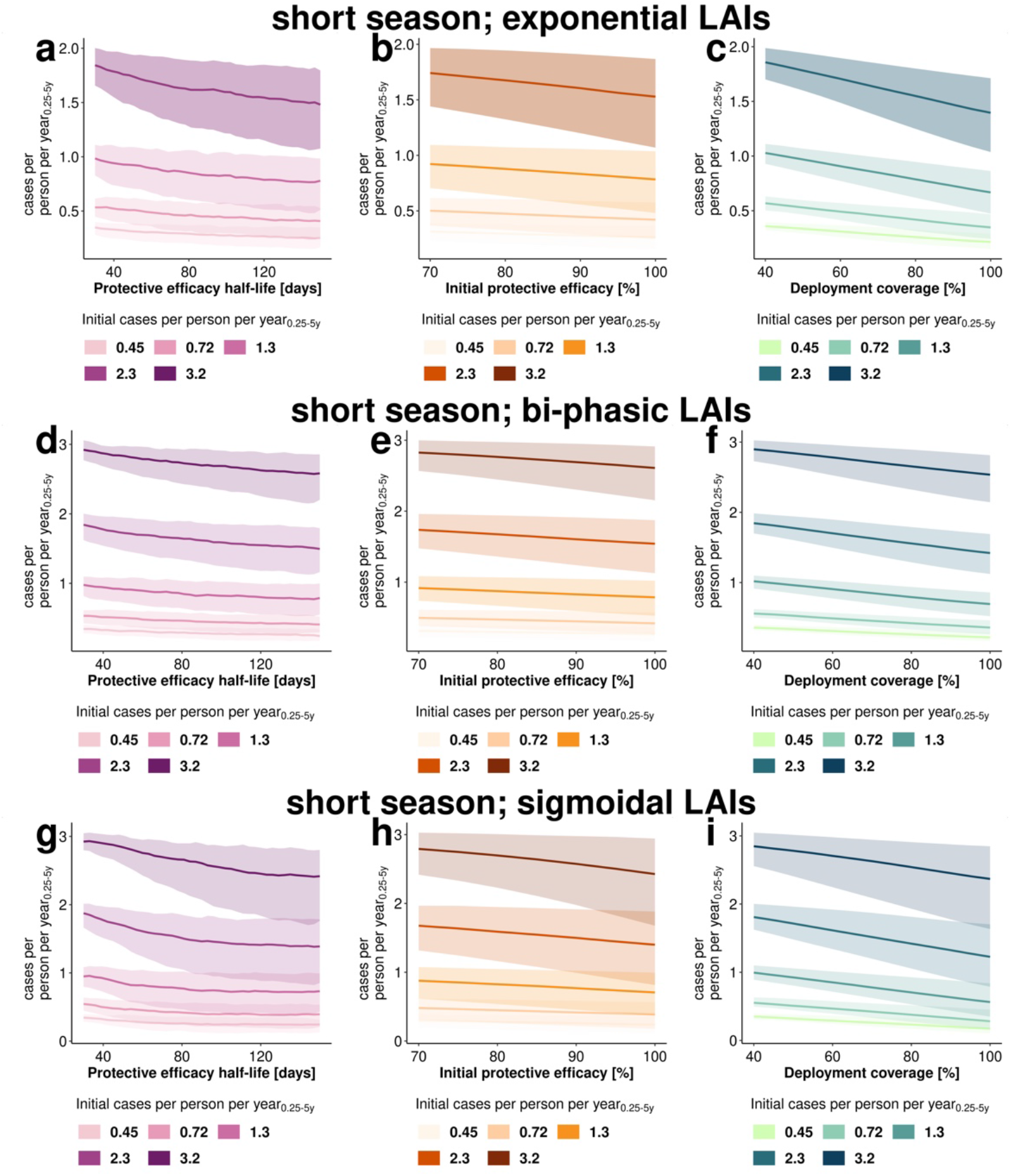
Estimated impact of LAI properties and operational factors on the level of clinical incidence reduction. Results are shown for the implementation stage in a setting with low access to care (E_14_=0.1) and short (Senegal-like) malaria transmission season, for **(a-c)** *exponential LAIs*, **(d-f)** *bi-phasic LAIs*, and **(g-i)** *sigmoidal LAIs*. Changes in clinical incidence measured as cases per person per year_0.25-5y_ with increasing tool properties or deployment coverage across the parameter space are shown for for **(a, d, g)** half-life (30-150 d), **(b, e, h)** initial efficacy (80-100%) and **(c, f, i)** coverage (40 - 100 %). The lines represent the mean and the 95%- confidence bands (shaded area) capture the distribution of incidence reduction across all sampled values. Increasing color intensity represents increasing initial cases per person per year_0.25-5y_). These results hold true for high access to healthcare settings. The conversion of initial cases per person per year_0.25-5y_ to prevalence can be found in Table S1.

**Figure S 6:**
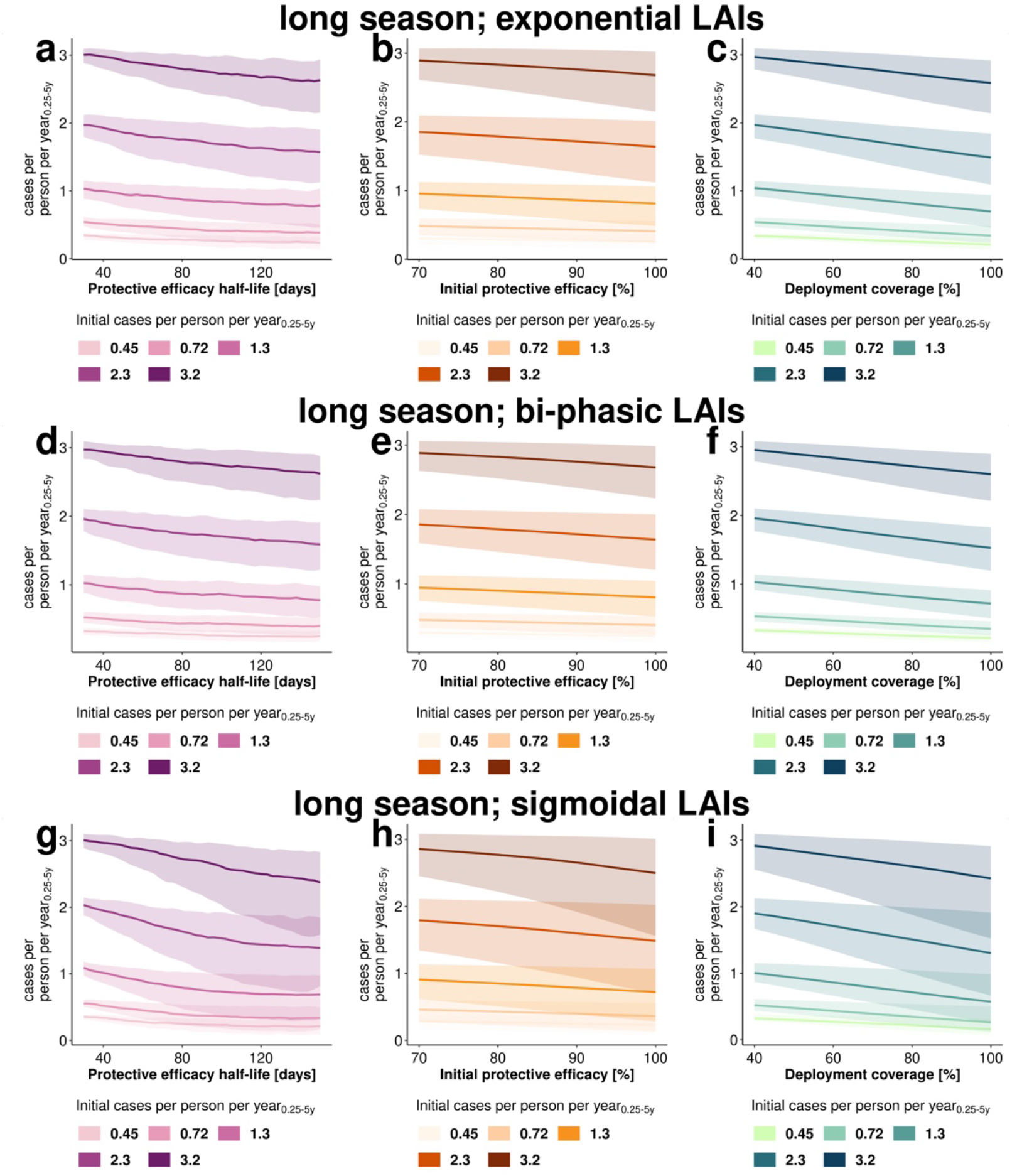
Estimated impact of LAI properties and operational factors on the level of clinical incidence reduction. Results are shown for the implementation stage in a setting with low access to care (E_14_=0.1) and long (Mali-like) malaria transmission season, for **(a-c)** *exponential LAIs*, **(d-f)** *bi-phasic LAIs*, and **(g-i)** *sigmoidal LAIs*. Changes in clinical incidence measured as cases per person per year_0.25-5y_ with increasing tool properties or deployment coverage across the parameter space are shown for for **(a, d, g)** half-life (30-150 d), **(b, e, h)** initial efficacy (80-100%) and **(c, f, i)** coverage (40 -100 %). The lines represent the mean and the 95%- confidence bands (shaded area) capture the distribution of incidence reduction across all sampled values. Increasing color intensity represents increasing initial clinical incidence (cases per person per year_0.25-5y_). These results hold true for high access to healthcare settings. The conversion of initial cases per person per year_0.25-5y_ to prevalence can be found in Table S1.

**Figure S 7:**
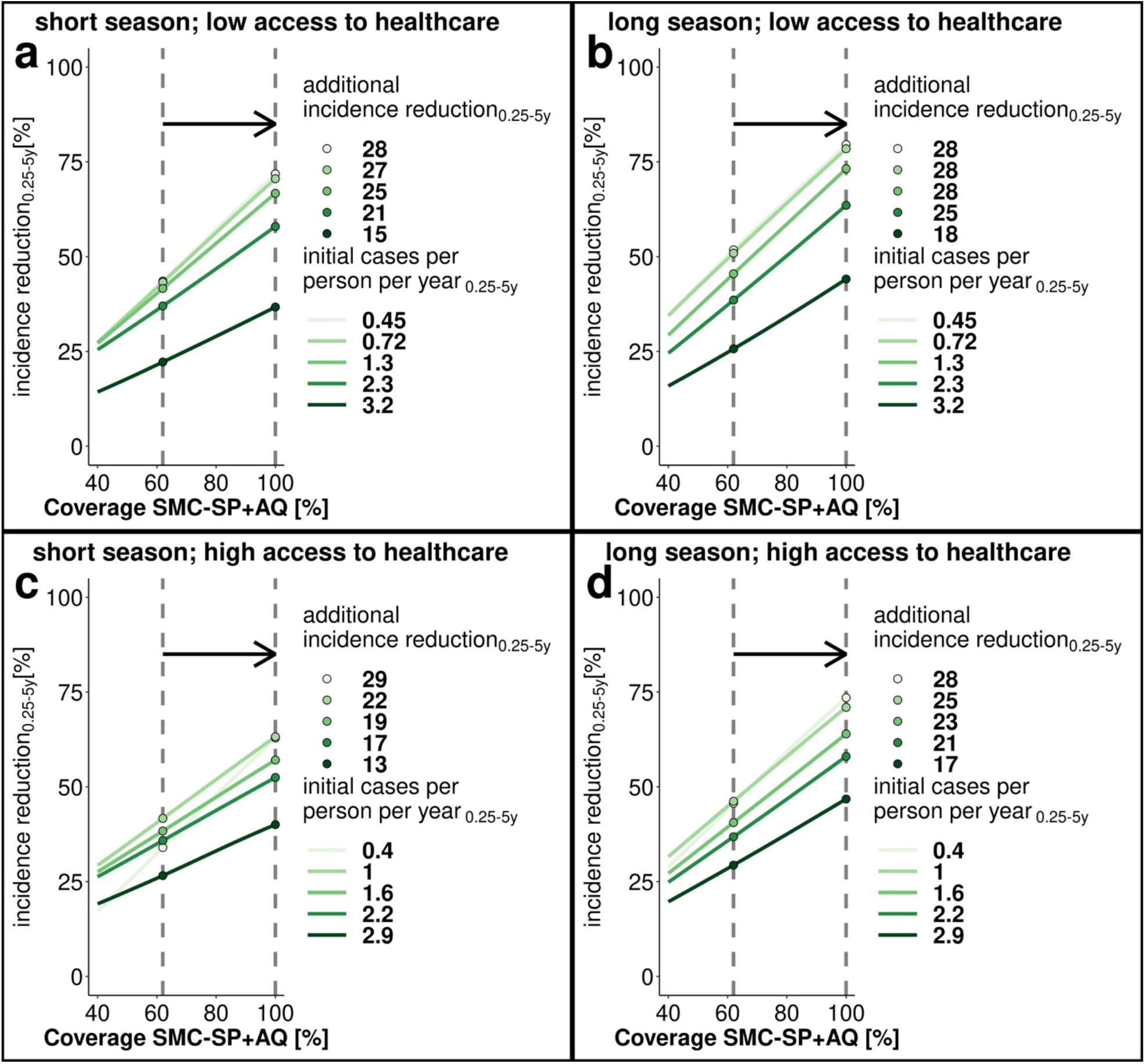
Incidence reduction achieved through implementation of SMC-SP+AQ over varying deployment coverage. The results are shown for low **(a, b)** and high **(c, d)** access to health care and short **(a, c)** and long **(b, d)** malaria seasons. The colors indicate different initial clinical incidence before introduction of SMC-SP+AQ (initial cases per person per year_0.25-5y_). The incidence reduction_0.25-5y_ was calculated in the implementation scenario after one year of implementation. The grey lines indicate the additional incidence reduction _0.25-5y_) achieved by increasing SMC-SP+AQ coverage from 62% to 100 %.

**Figure S 8:**
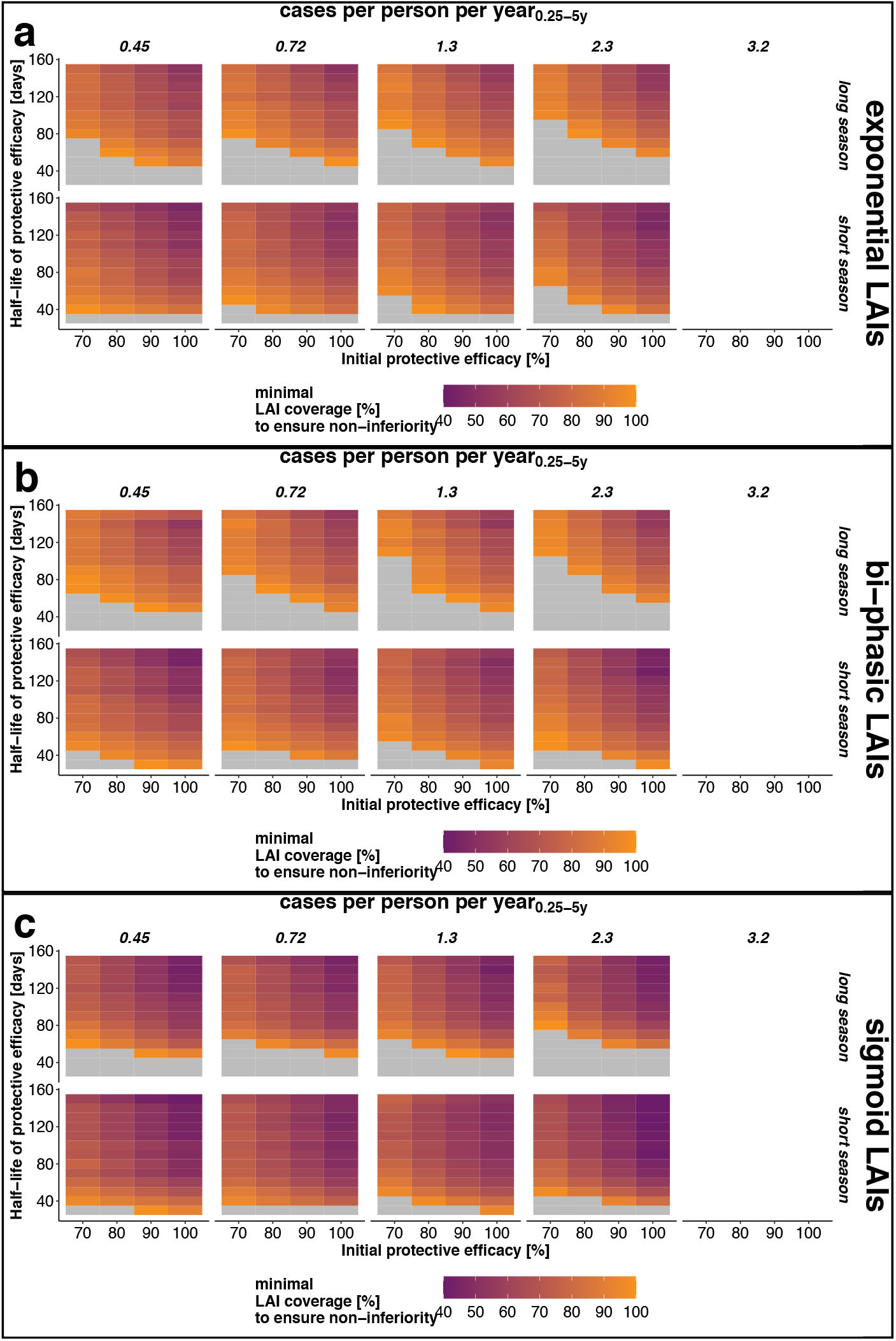
Estimated minimal LAI coverage required during *implementation stages* to achieve non-inferiority in a given setting. Heatmap of the estimated minimal coverage (colour) of LAIs at which non-inferiority to SMC-SP+AQ (assuming a fixed SMC coverage of 60% in each of the 3 or 4 rounds) is achieved for different combinations of protective efficacy decay, initial protective efficacy and protective efficacy half-life. The results are displayed for intervention scenarios with a low access to care (E_14_=0.1) in the two seasonal settings. In the grey area, non-inferiority could not be established for any combination of tool properties. The LAI coverage could not be optimized for high transmission settings (initial cases per person per year_0.25-5y_=3.2) because they fail to sufficiently protect the targeted population from clinical malaria even at full deployment coverage. Therefore, optimisation of the LAI deployment coverage could not be conducted (Fig. S4).

**Figure S9:**
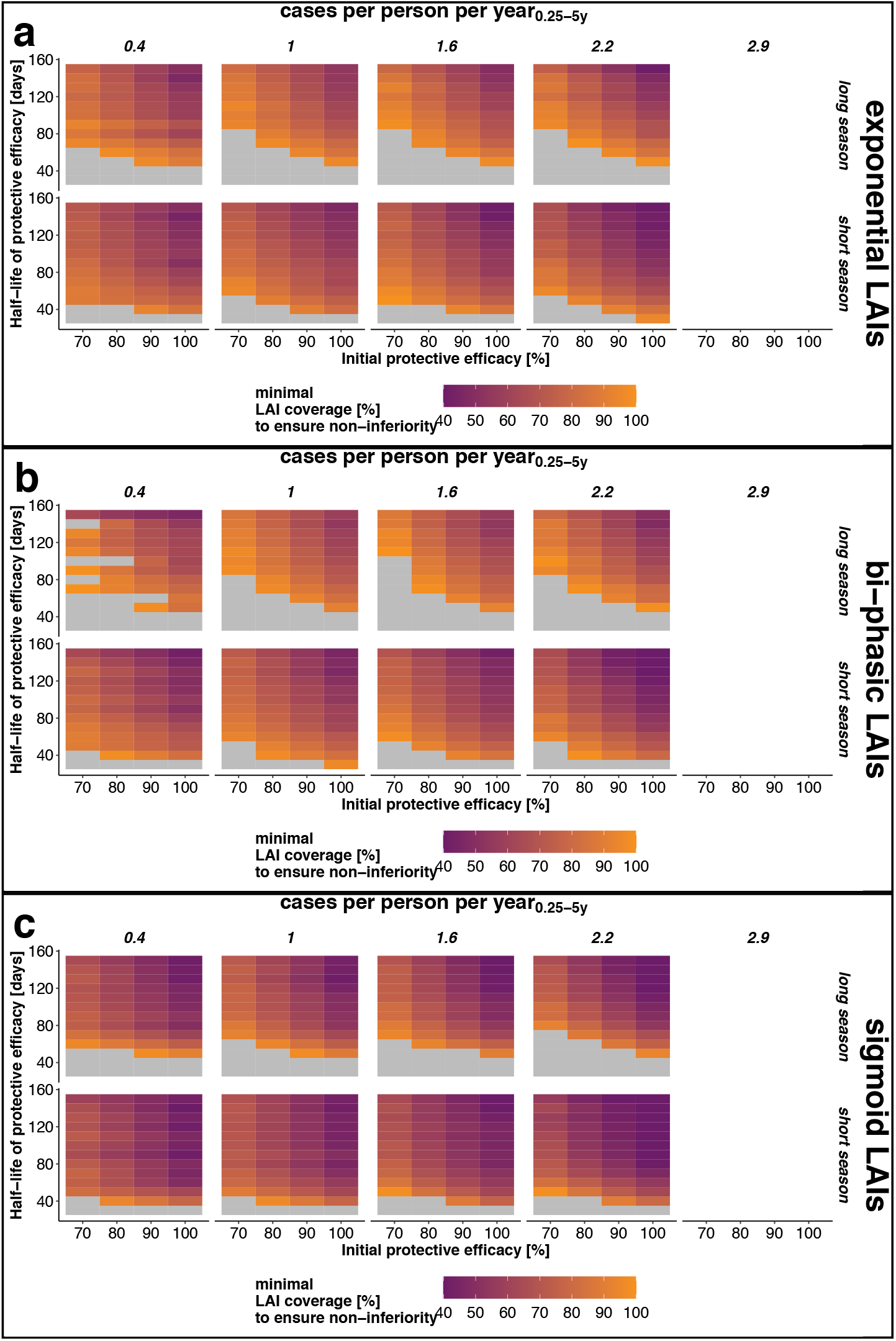
Estimated minimal LAI coverage required during *implementation stages* to achieve non-inferiority in a given setting. Heatmap of the estimated minimal coverage (colour) of LAIs at which non-inferiority to SMC-SP+AQ (assuming a fixed SMC coverage of 60% in each of the 3 or 4 rounds) is achieved for different combinations of protective efficacy decay, initial protective efficacy and protective efficacy half-life. The results are displayed for intervention scenarios with a high access to care (E_14_=0.5) in the two seasonal settings. In the grey area, non-inferiority could not be established for any combination of tool properties. The LAI coverage could not be optimized for high transmission settings (initial cases per person per year_0.25-5y_=2.9) because they fail to sufficiently protect the targeted population from clinical malaria even at full deployment coverage. Therefore, optimisation of the LAI deployment coverage could not be conducted (Fig. S4).

**Figure S10:**
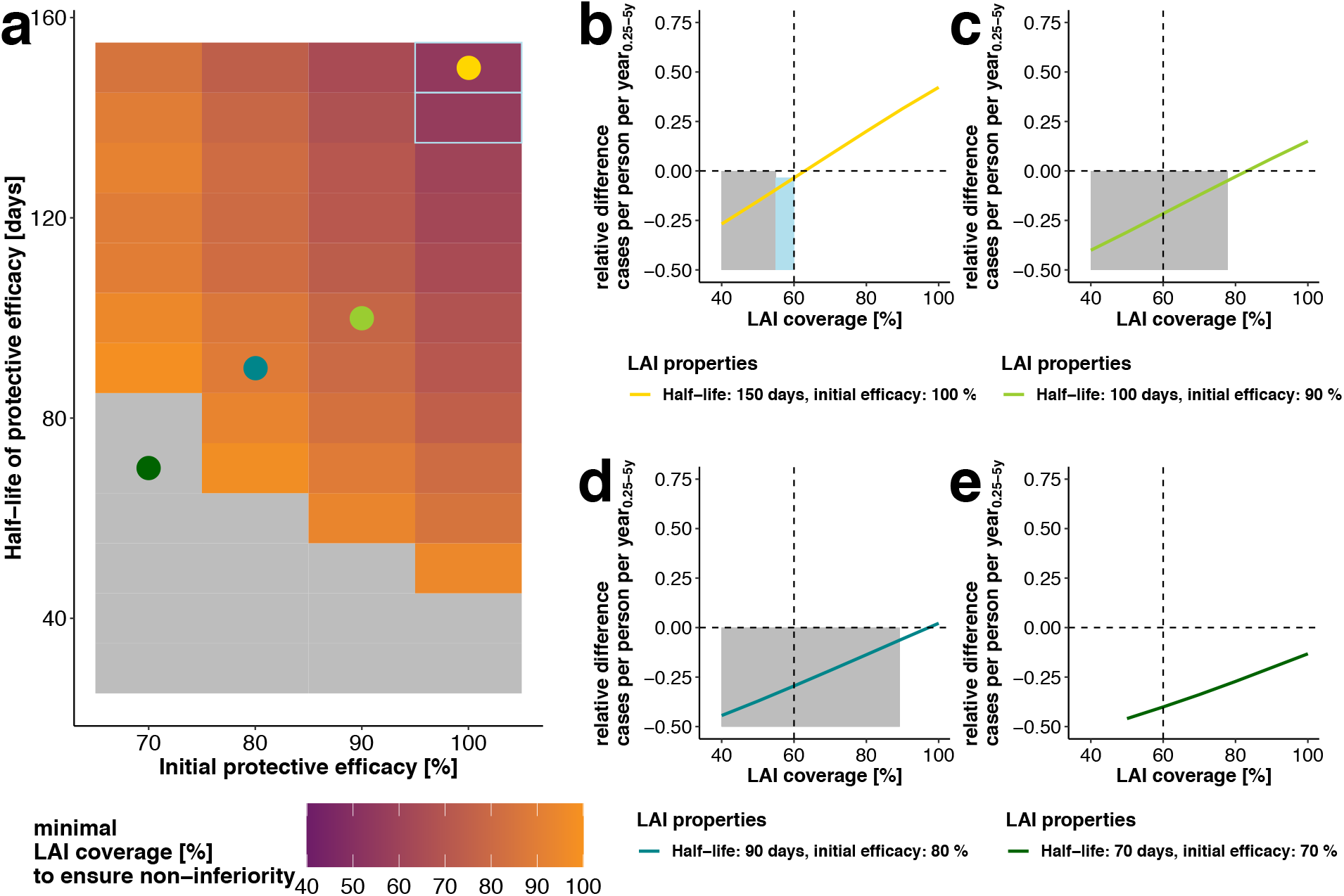
Estimated minimal LAI coverage required during *implementation stages* to achieve non-inferiority in a given setting and predicted gains in cases averted of subsequent *exponential* LAI coverage increments. (**(a)** Heatmap of the estimated minimal coverage (colour) of *sigmoidal LAIs* at which non-inferiority to SMC-SP+AQ (assuming a fixed SMC coverage of 60%) is achieved for different combinations of *exponential LAI* efficacy and half-life. The results are displayed for intervention scenarios with an underlying disease burden of 1.4 cases per person per year_0.25-5y_, long malaria transmission season and low access to treatment (E_14_=0.1). In the grey area, non-inferiority could not be established for any coverage. For the tool characteristics within the light-blue frames, non-inferiority could be reached with a LAI coverage under the reference SMC-SP+AQ coverage of 60%. The coloured dots represent four illustrative LAI profiles of **(b)**150 days half-life and 100% initial efficacy (yellow), **(c)** 100 days half-life and 90% initial efficacy (light green), **(d)** 90 days half-life and 80% initial efficacy (blue), and **(e)** 70 days half-life and 70% initial efficacy (dark green, e). **(b-e)**. Corresponding predicted relative differences in cases per person per year_0.25-5y_ (Eq. 5) are calculated for the illustrative LAIs (coloured dots) in (a) in the last implementation year (5 years after LAI introduction) over all LAI coverages as compared to SMC-SP+AQ at 60% coverage (vertical dotted line). The predicted positive increase in relative difference in yearly clinical cases (above the dotted horizontal line) means more clinical cases are averted with LAIs than with SMC-SP+AQ. It thus illustrates the benefit of increasing *exponential LAI*-coverage above the minimal required coverage to achieve non-inferiority (shown by the grey coloured area). In the light-blue area in (b), a LAI coverage lower than the SMC-SP+AQ coverage is sufficient to establish non-inferiority.

## 6. Parameterization of SMC-SP+AQ to clinical trial data

Seasonal malaria chemoprotection (SMC) was implemented as sulfadoxine–pyrimethamine+amodiquine (SP-AQ) treatment and calibrated to a randomized non-inferiority trial of dihydroartemisinin-piperaquine with SP-AQ, conducted between August 2009 and January 2010 in rural western Burkina Faso Zongo et al.(2015)^3^. Decay of protective efficacy of SPAQ in the field over time was extracted from Fig.3 in Zongo et al.(2015)^3^ and used to parameterized the decay functions as specified in *OpenMalaria* using a least squares approach combined with a Gaussian-Process optimization. The trial described in Zongo et al.(2015)^1^ was rebuilt in OM settings listed in Table S4.

**Table S 4:**
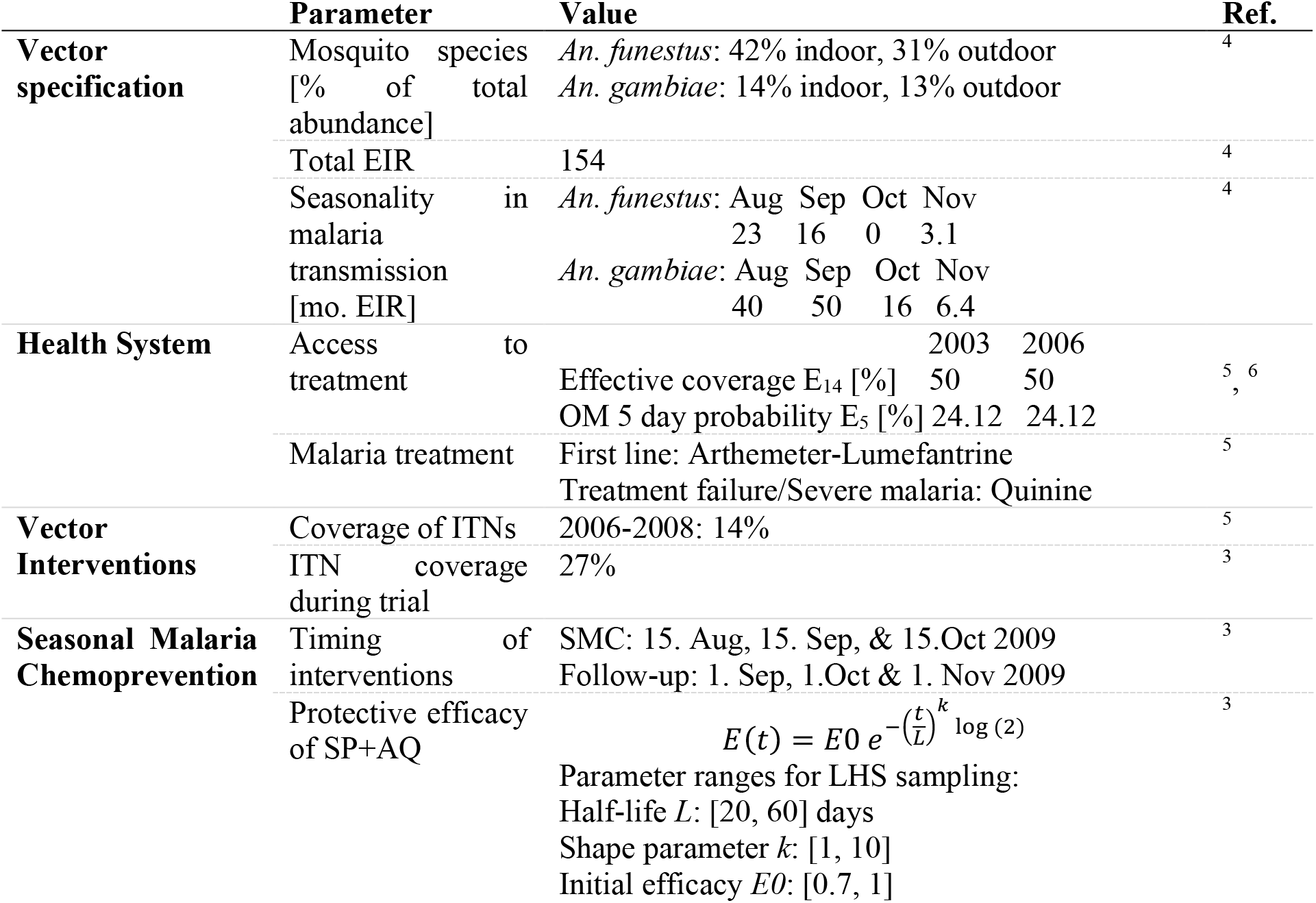
Inputs into OpenMalaria to check adequate parameterization of SMC with SP-AQ.

We used Latin-Hypercube sampling (LHS) to generate 5000 samples of the decay function parameters for protective efficacy (L, k, E0) within the parameter bounds given in Table S4. The trial was simulated with these parameters and five seeds per parameter-set for the intervention and control cohort. The protective efficacy *E* of SMC -SP+AQ in the simulated trial compared to the simulated controls (without intervention) was extracted by comparing cases per person (cpp) between the intervention group (cpp_int_) and control group (cpp_cont_) over the trial period as follows:

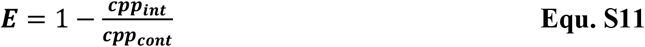

The residual sum of squares (RSS) between the protective efficacy given in Zongo et al.(2015)^3^ and protective efficacy from OM simulations was extracted and a Gaussian process (GP) regression was trained to predict the RSS between trial results and OM simulation with the parameters of the protective efficacy decay.

The true over predicted RSS of the hold-out of 1000 data points is shown in Fig. S11.

**Figure S 11:**
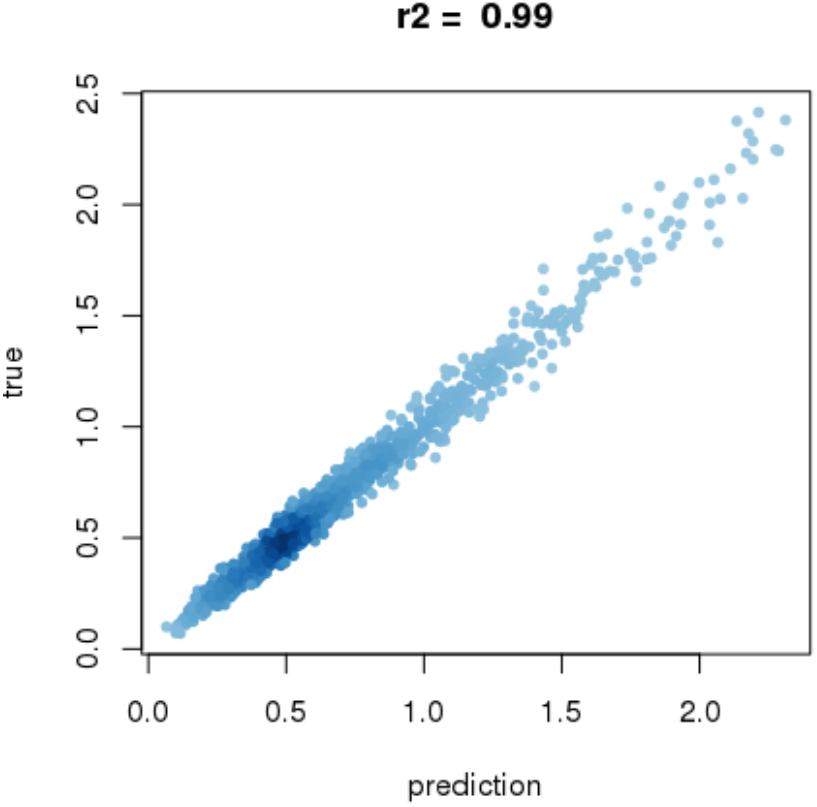
True over predicted RSS of the GP.

The GP was then optimized via non-linear optimization using the augmented Lagrange method (function gosolnp, R-package Rsolnp). We optimized the mean predicted error +/- one or two standard deviation of the residual sum of squares (RSS) between the protective efficacy in Zongo et al.(2015)^3^ and the OM simulation output. The parameters of the decay function returned by the optimization process were re-simulated with OM and the RSS extracted (*RSS OM*). The OM simulation resulting in the least RSS was then selected for further analysis.

**Table S 5:** Results of the GP optimization. The results of the optimization *RSS optim* is compared with the *predicted (pred*.*) RSS* by the GP and the RSS extracted from OM simulations (median of 5 seeds) conducted with the respective parameters (*RSS OM*). The solver was restarted 20 times and the number of random parameters generated for every restart was set to n.sim=500.

|  | kdecay | Half-life | Initial efficacy | RSS optim | RSS pred | RSS OM |
| --- | --- | --- | --- | --- | --- | --- |
| **Min. RSS before optimisation** | 6.35 | 32.5 | 0.97 | - | 0.0842 | 0.0601 |
| **mean** | 5.40 | 31.3 | 1.0 | 0.0538 | 0.0538 | **0.0556** |
| **Mean +sd** | 5.29 | 31.5 | 1.0 | 0.0882 | 0.0542 | 0.0583 |
| **Mean - sd** | 5.58 | 31.0 | 1.0 | 0.0186 | 0.0545 | 0.0538 |
| **Mean + 2\*sd** | 5.28 | 31.5 | 1.0 | 0.0895 | 0.0541 | 0.0556 |
| **Mean - 2\*sd** | 5.62 | 31.1 | 1.0 | 0.0172 | 0.0544 | 0.0739 |

The OM implementation of the best parameter set for the intervention cohort is able to well capture the protective efficacy described in ^3^. The protective efficacy (Figure S12), cumulative hazard (Fig. S13) and prevalence over time (Fig. S14) provide more insight into the trial results and comparison with OM outputs.

**Table S 6:** Comparison of trial results of Zongo et al., 2015 ^3^ and model outputs using the model specification in Table S4 with the best parameter set. The compared time-points correspond to the time-points compared in ^**3**^. The difference in prevalence of parasitemia in December 2009 in the untreated cohort are caused by ongoing but decreasing transmission in December (see Figure S3) and unclear definition of data time-point in ^3^.

|  | Zongo et al., 2015 | OM<br>[min, max] |
| --- | --- | --- |
| Prevalence of parasitemia [%] in August 2009, intervention cohort | 45 | 45<br>[44, 48] |
| Prevalence of parasitemia [%] in September 2009, untreated cohort | 61 | 60<br>[57, 65] |
| Cumulative hazard for fever with any parasitemia intervention cohort | 0.22 | 0.14<br>[0.13, 0.15] |
| Cumulative hazard for fever with any parasitemia intervention cohort | 0.92 | 1.01<br>[0.93, 1.12] |
| Prevalence of parasitemia [%] in November 2009, intervention cohort | 12 | 19<br>[18, 20] |
| Prevalence of parasitemia [%] in December 2009, untreated cohort | 36 | 46<br>[39, 48] |

**Figure S 12:**
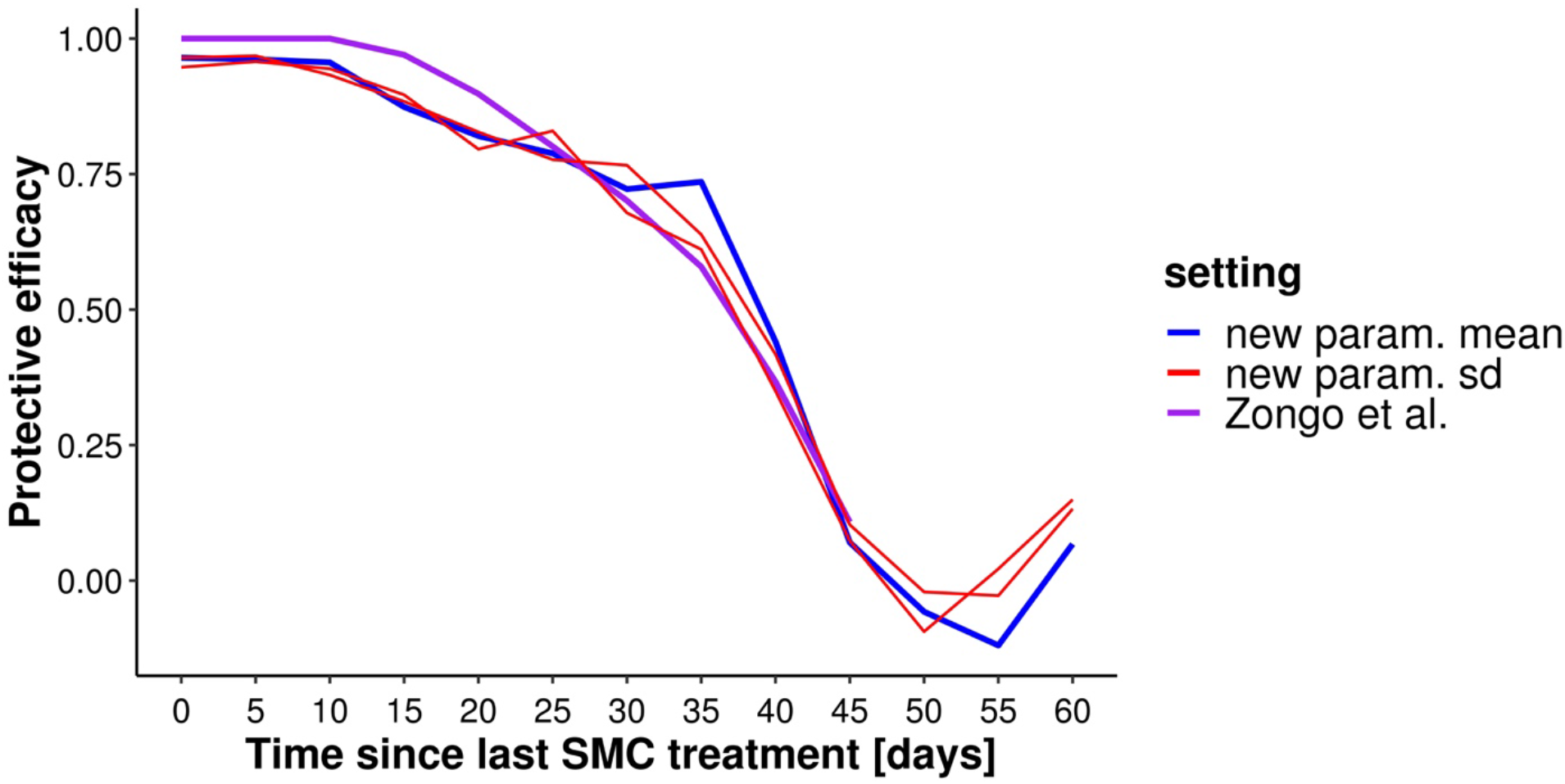
Decay of protective efficacy of SMC-SP+AQ over time. Population protective efficacy over time in the trial setting. The purple line represents the protective efficacy as extracted from Zongo et al.(2015) ^**3**^. The new parameterization after optimization and uncertainty are shown in blue and red.

**Figure S 13:**
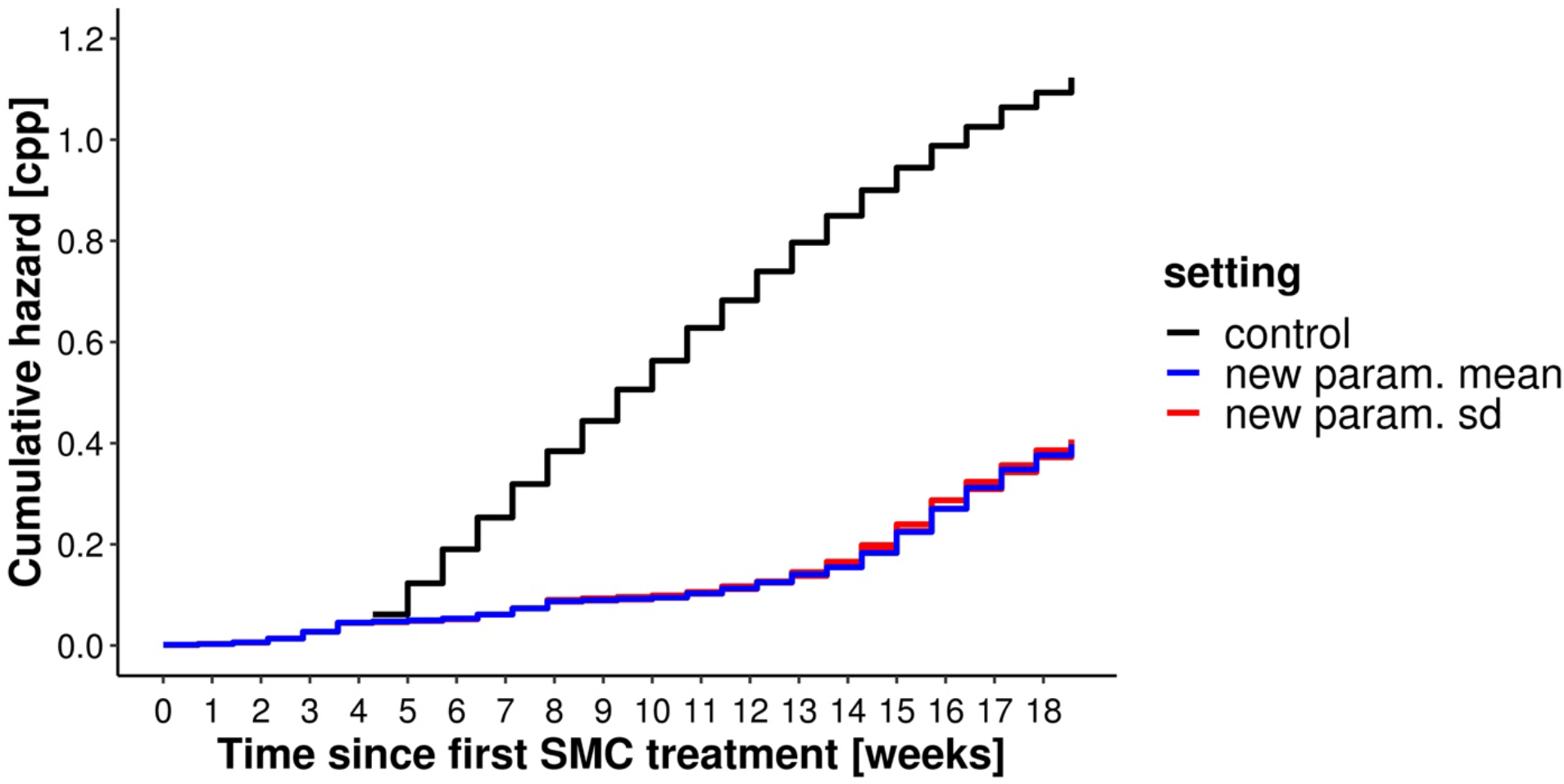
Cumulative hazard of malaria in children who received SMC with SP-AQ (blue) compared to controls (black). The cumulative hazard over time was calculated using the Nelson-Aalen estimator. See Fig. 2 in Zongo et al.(2015)^**3**^ for comparison.

**Figure S 14:**
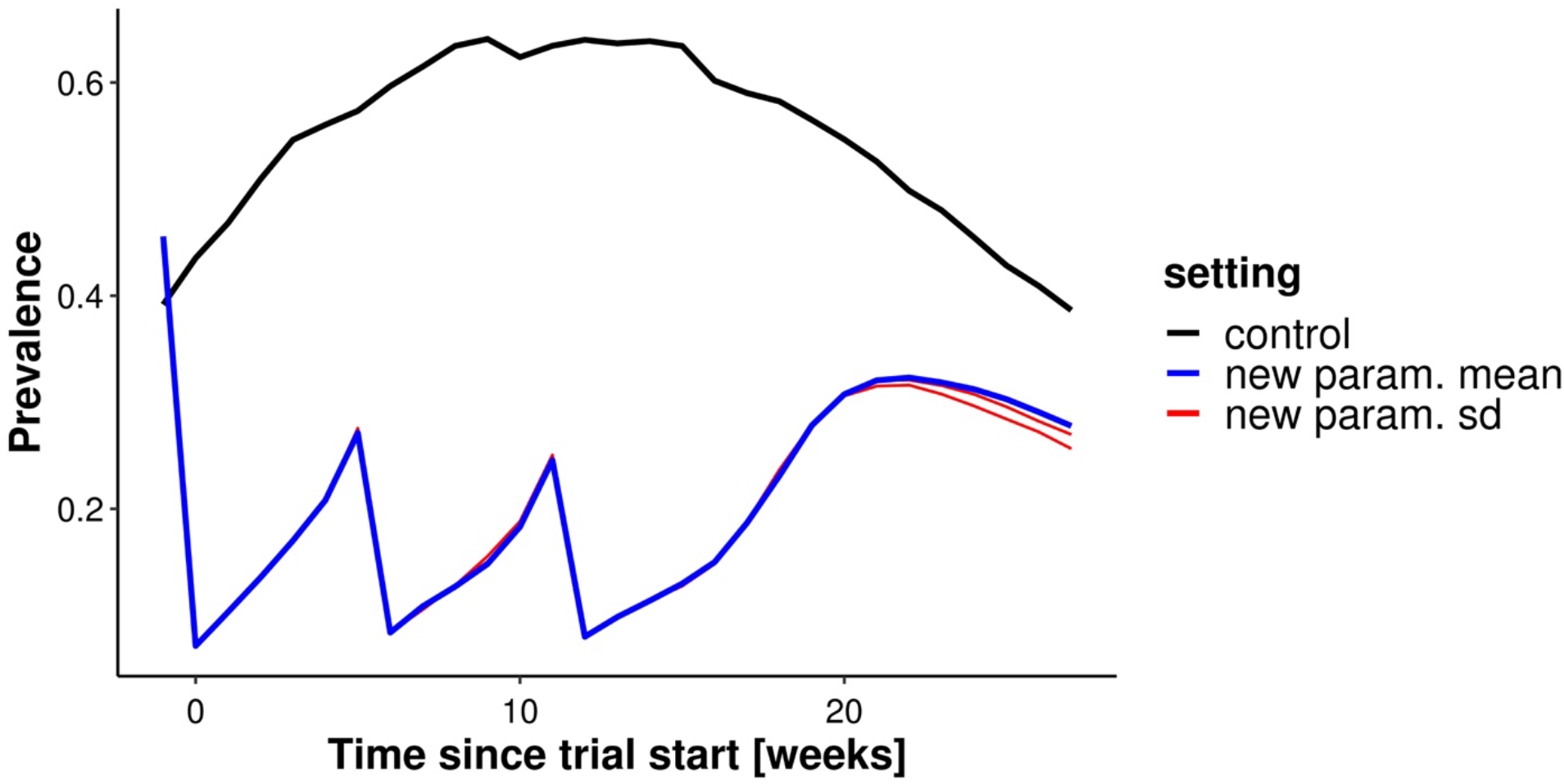
Prevalence of malaria in children who received SMC with SP-AQ (blue) compared to controls (black) over the trial. The prevalence in the intervention cohort is decreasing with each SMC round every month and then slowly increasing again. At the same time, the prevalence in the control group is increasing with the ongoing malaria transmission season. After the last SMC round, there is an increase of prevalence caused by still ongoing transmission. Simultaneously, the transmission intensity is already decreasing as can be seen in the control cohort prevalence.

## Notes

### Clinical Trial

Not applicable

### Author Declarations

As this is a simulation study no IRB or ethics committee approvals are required

### Summary of Updates

We updated funding and role of funders

